# Circulating immunome fingerprint in eosinophilic esophagitis is associated with clinical response to proton pump inhibitor treatment

**DOI:** 10.1101/2023.12.29.23300619

**Authors:** Lola Ugalde-Triviño, Francisca Molina-Jiménez, Juan H-Vázquez, Carlos Relaño-Rupérez, Laura Arias-González, Sergio Casabona, María Teresa Pérez-Fernández, Verónica Martín-Domínguez, Jennifer Fernández-Pacheco, Alfredo J Lucendo, David Bernardo, Cecilio Santander, Pedro Majano

**Affiliations:** Molecular Biology Unit, Hospital Universitario de la Princesa, Madrid, Spain; Instituto de Investigación Sanitaria Hospital Universitario de La Princesa (IIS-Princesa), Madrid, Spain; Mucosal Immunology Lab, Centro de Investigaciones Biomédicas en Red de Enfermedades Infecciosas (CIBERINFEC), Unidad de Excelencia Instituto de Biología y Genética Molecular (IBGM), Universidad de Valladolid, 47005 Valladolid, Spain; Bioinformatics Unit, Centro Nacional de Investigaciones Cardiovasculares (CNIC), Madrid, Spain; Centro de Investigación Biomédica en Red de Enfermedades Infecciosas (CIBERINFEC), 28029 Madrid, Spain; Department of Gastroenterology, Hospital General de Tomelloso, Tomelloso, Ciudad Real, Spain; Instituto de Investigación Sanitaria de Castilla-La Mancha (IDISCAM), Spain; Centro de Investigación Biomédica en Red de Enfermedades Hepáticas y Digestivas (CIBERehd), Madrid, Spain; Department of Gastroenterology, Hospital Universitario de La Princesa, Madrid, Spain; Department of Cellular Biology, Faculty of Biology, Universidad Complutense de Madrid, Madrid, Spain

**Keywords:** Spectral cytometry, Biomarker, Eosinophilic esophagitis, Plasmacytoid dendritic cells

## Abstract

**Objectives:** The aim of the study was to characterize the circulating immunome of patients with EoE before and after proton pump inhibitor (PPI) treatment in order to identify potential non-invasive biomarkers of treatment response.

**Methods:** PBMCs from 19 healthy controls and 24 EoE patients were studied using a 39-plex spectral cytometry panel. The plasmacytoid dendritic cell (pDC) population was differentially characterized by spectral cytometry analysis of immunofluorescence assays in esophageal biopsies from 7 healthy controls and 13 EoE patients.

**Results:** Interestingly, EoE patients at baseline had lower levels of circulating pDC compared with controls. Before treatment, patients with EoE who responded to PPI therapy had higher levels of circulating pDC and classical monocytes, compared with non-responders. Moreover, following PPI therapy pDC levels were increased in all EoE patients, while normal levels were only restored in PPI-responding patients. Finally, circulating pDC levels inversely correlated with peak eosinophil count and pDC count in esophageal biopsies. The number of tissue pDCs significantly increased during active EoE, being even higher in non-responder patients when compared to responder patients pre-PPI. pDC levels decreased after PPI intake, being further restored almost to control levels in responder patients post-PPI.

**Conclusions:** We hereby describe a unique immune fingerprint of EoE patients at diagnosis. Moreover, circulating pDC may be also used as a novel non-invasive biomarker to predict subsequent response to PPI treatment.

**WHAT IS KNOWN:**

- Eosinophilic esophagitis (EoE) is a Th2 type immune disorder with increased prevalence in the last years.
- Proton pump inhibitor (PPI) treatment is the preferred first-line therapy for EoE, which leads to clinical and pathological reversion in about half of the cases. Non-responding patients require other therapeutic options.
- Currently, an endoscopy with esophageal biopsies is required for EoE diagnosis and to monitor response to treatment. Therefore, the discovery of non-invasive biomarkers is of utmost importance for treatment monitoring.
- To date only peripheral eosinophil and Th2 profiles have been studied in EoE.

**WHAT IS NEW HERE:**

- At diagnosis EoE patients have a specific circulating immune signature.
- PPI-responding and non-responding EoE patients have different immune fingerprints at baseline.
- Immune characterization of EoE patients at diagnosis and after PPI treatment unveiled differential levels of circulating plasmacytoid dendritic cells (pDCs) depending on their inflammatory state and response to PPI treatment. Those levels are related with the number of pDCs infiltrated in the esophageal tissue.

## INTRODUCTION

Eosinophilic esophagitis (EoE) is a Th2-type immune disorder which is considered an increasing leading cause of chronic esophageal dysfunction in patients of all ages^1,2^ just after gastro-esophageal reflux disease (GERD).

Eosinophils are normally found in the gastrointestinal tract; however, they are absent from the esophageal tissue in health conditions. In EoE, the eosinophil infiltrate in the esophageal tissue layers^3,4^ leads to tissue remodelling and fibrosis as well as subsequent dysfunction characterized by esophageal dysmotility, narrowing and rigidity^5^. As a result, patients experience food impaction, dysphagia and heartburn among other symptoms, which impair their health-related quality of life. Therefore, an early EoE diagnosis and effective therapy are essential to prevent impairment of esophageal function. EoE patients often have concurrent allergic responses to food and airborne allergens, together with a yet unexplained male predominance^6,7^.

First-line therapeutic options for EoE include dietary restrictions, protein pump inhibitor (PPI) therapy and swallowed topical corticosteroids, which provide variable effectiveness^8–12^. Recently, the anti-interleukin-4 receptor antagonist dupilumab joined the therapeutic armamentarium against EoE^13,14^. Among them, PPI represent the preferred therapy in clinical practice, despite its limited effectiveness^15^; histologic remission and clinical improvement after PPI are achieved by only 50% and 70% of treated patients, respectively^16^ . Patients who do not respond to PPI require subsequent therapeutic options.

Currently, EoE diagnosis and treatment response monitoring require endoscopy with esophageal biopsies, as clinical symptoms do not correlate well with esophageal inflammation^17,18^. In this regard, the chronic nature of this disease together with the dissociation between patients’ symptoms and esophageal inflammation^18^ require seeking for novel reliable biomarkers and clinical parameters able to identify patient profiles at diagnosis and follow-up. Recent studies have focused on seeking for new biomarkers by characterizing the RNA^19–21^ and proteomic profile of EoE at esophageal tissue level^22^. Nevertheless, these approaches are still invasive. Hence, the development of novel non-invasive biomarkers to aid on EoE diagnosis and monitoring is an essential unmet need^23^.

Building from all these precedents, in this study we aimed to characterize the circulating immunome of EoE patients at the time of diagnosis using top-of-the-art spectral cytometry. With this approach, we expected to identify specific immune subsets that could not only help to characterize this disorder, but also to identify novel non-invasive biomarkers able to predict response to PPI treatment.

## MATERIAL AND METHODS

### Human subjects

A total of 25 incident adult EoE patients were prospectively recruited at the moment of diagnosis at Hospital Universitario de La Princesa (Madrid, Spain) between February 2018 and November 2020. EoE was diagnosed according to evidence-based guidelines^3^ including: (i) symptoms referring to esophageal dysfunction, (ii) infiltration of the esophageal epithelium by 15 or more eosinophils per high-powered field (hpf) assessed from 6 endoscopic biopsy samples; (ii) absence of eosinophilic infiltration in biopsy specimens from gastric and duodenal mucosa; and (iii) exclusion of alternative potential causes of esophageal eosinophilia. Subjects (n=19) who underwent upper endoscopy for assessment of dyspepsia or suspected gastroduodenal ulcer were included as controls. Esophageal biopsies obtained in the endoscopy were normal in all cases and eosinophilic infiltration was excluded. All controls with hiatus hernia, incompetent cardias, or esophageal peptic lesions were excluded hence ensuring that all the recruited controls displayed a normal endoscopic appearance and eosinophil-free biopsies of the esophagus.

Atopic background was recorded for all EoE patients and control subjects. The EoE endoscopic reference score (EREFS) rating the severity of esophageal inflammation (oedema, furrows, exudates) and fibrosis (rings and stricture)^24^ was assessed in all patients. Furthermore, the validated Eosinophilic Esophagitis Histologic Scoring System (EoEHSS) was also determined, evaluating eight pathologic features for both severity (grade) and extent (stage) of abnormalities^25^.

Out of the 25 EoE patients, 18 underwent double dose PPI treatment (omeprazole 20 mg b.i.d. or equivalent) for an 8-week period, after which they were classified as responders (n=9) or non-responders (n=9) based on the peak eosinophil count in a second endoscopy in which EREF and EoEHSS scores were also assessed. The study was approved by the local Ethics Committee (PI17/0008, registry number 3107, 8 June 2017). All individuals provided written informed consent.

### Blood samples

In all cases, blood samples were obtained at the time of the endoscopy from EoE patients (both before and after PPI-treatment in the latter) and controls. After placing a venous line to provide sedation for endoscopy, blood was collected in EDTA-coated tubes to isolate peripheral blood mononuclear cells (PBMC).

### PBMCs staining and spectral cytometry acquisition

PBMCs were isolated by Ficoll gradient assay. Viable cells were counted and cryopreserved in freezing medium (Foetal bovine serum [Hyclone,Thermofisher] complemented with DMSO 20% medium) at vapour phase of liquid nitrogen.

For the analysis, PBMCs were thawed and a total of 2 million PBMCs were stained with monoclonal antibodies (Supplementary Table 1) applying a modified OMIP-69 panel protocol^26^. Before staining, Live/Dead fixable blue dead cell stain kit was added to exclude dead cells from the analysis. Brilliant Stain Buffer and True-Stain Monocyte Blocker were also added before staining with the aim of obtaining the optimal fluorescence. PBMCs were washed with FACS buffer (500mL PBS +1 0mL filtered FCS + 0.1g NaN_3_ + 2.5mL sterile EDTA) and incubated in the dark at room temperature during staining. Cells were further fixed in 0.8% paraformaldehyde in FACS buffer in the dark for 10min and washed with FACs buffer. Cells were preserved at 4°C until acquired (within 48h) in a 5-laser spectral cytometer (Aurora, Cytek).

### Cytometry data and statistical analysis

Spectral cytometry data were analysed using the OMIQ Data Science platform (© Omiq, Inc. 2022). After setting the scale, parameters, and cofactors, the FlowAI algorithm was used for cleaning the data from aberrant signal patterns or events. Then, cell debris and doublets were excluded to gate viable leukocytes (CD45^+^) where an unsupervised approach was applied with a dimensionality reduction Uniform Manifold Approximation and Projection (UMAP)^27^ plus clustering FlowSOM algorithms. Merging these two algorithms allows a deeper classification of the different immune subsets through different marker expression on the UMAP. A heatmap was also built showing the expression levels of each marker within each cluster. Dendrograms further grouped the clusters and markers associated by similarity.

Statistical analysis was performed in all cases using Rstudio 2022.07.2+576. Differential analysis of clusters defined in OMIQ was performed with the edgeR package, using genewise quasi-likelihood (QL) F-tests with GLMs. Significance was set at p-value≤ 0.05 and Log2 Fold Change (LogFC) ≥1.5. Differences between groups of significant clusters were validated by classic gating strategy approaches (Supplementary Figure 1 and 2). Manual gating results were analysed by *t*-test (paired when indicated) and/or two-way ANOVA test followed by *post hoc* Fisher test. Outliers were determined through Grubbs’ test and deleted from the final analysis. Statistical significance was considered when p-value ≤0.05 in all cases (p< 0.05 *, p< 0.01 **, p<0.001 ***, p<0.0001****, n.s.= not significant). Percent of total always refers to percentage of cells of the specified population relative to total PBMCs.

### Immunofluorescence staining in esophageal biopsies

Immunofluorescence staining was performed in biopsies from control subjects and EoE patients, previously oriented in a cellulose acetate and included in paraffin-embedded blocks, using the antibodies CD123-biotin (Stem Cell, 60110BT), HLA-DR (Santa Cruz, sc-53319) and CD11c (Thermo Fisher Scientific, PA535326). Briefly, slides with 4-μm sections of esophageal biopsies, underwent deparaffinization, antigen retrieval using sodium citrate buffer (pH 6.0), blocked with 4% goat serum/phosphate-buffered saline (PBS), and then incubated with primary antibody (HLA-DR and CD11c) diluted in 1% goat serum-PBS overnight at 4 °C in a humidified chamber. Then, slides were washed with PBS + 1% NP-40 and incubated with secondary antibodies diluted in 1% goat serum-PBS for 30min at RT in a humidified chamber. After, slides were washed with PBS + 1% NP-40 and incubated with primary antibody (CD123) diluted in 1% goat serum/PBS overnight at 4 °C in a humidified chamber. Finally, slides were washed in PBS + 1% NP-40 and incubated with secondary antibodies and DAPI (0.5 μg/mL) diluted in 1% goat serum/PBS for 30min at RT in a humidified chamber. Finally, a cover slip was added with ProLong Gold mounting reagent (Molecular Probes). Images were obtained using a Thunder Imager (Leica) with LAS X software and analysis was done using ImageJ and RStudio software. Normality was assessed by Saphiro Wilk normality test, the non-parametric Mann-Whitney test was done to compare between pairs of groups. In all cases (p< 0.05 *, p< 0.01 **, p<0.001 ***, p<0.0001****, n.s.= not significant).

## RESULTS

### Patient Demographics

Clinical and demographic characteristics of study participants are summarized in Table 1. Compared to controls (n=19), EoE patients (n=25) were older (34 vs 41 years) and more frequently male (68% vs 88%), but no significant differences were found in demographic characteristics across groups.

EoE patients treated with PPI (n=18) were classified as responders or non-responders to the therapy. EoE remission was defined as presenting a peak eosinophil count below 15 after at least 8 weeks of treatment. Response to PPI was also associated with improvement in EoE symptoms from baseline (assessed as patient reported outcomes), as well as in endoscopic and histologic characteristics, assessed with the scores EREFS and Grade and Stage EoEHSS, respectively. Baseline characteristics of PPI responding, and non-responding patients showed no differences.

**Table 1.**
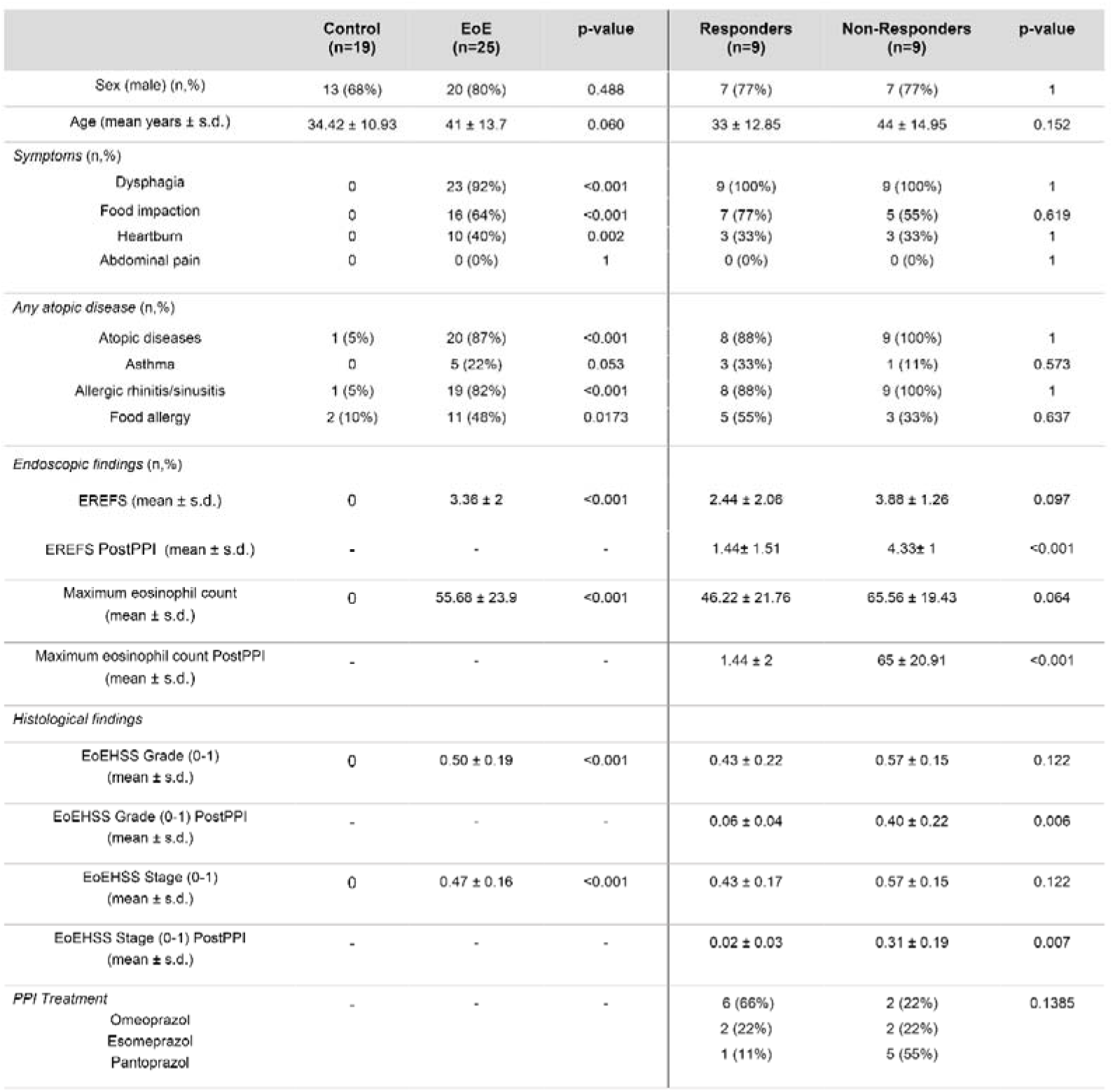
Clinical features of controls and EoE patients. Fisher’s test and *t*-test were applied to analyse differences between control and EoE patients and between responders and non-responders. P-value is shown for each comparison. Abbreviations: EoE: Eosinophilic Esophagitis; EoEHSS: Eosinophilic Esophagitis Histologic Scoring System; EREFS: EoE endoscopic reference score; PPI: Proton Pump Inhibitors.

### High dimensional analysis on PBMCs from controls and EoE patients

A total of 61 samples (19 controls, 24 patients at disease onset, as well as 9 responders and 9 non-responders to PPI after 8-week treatment) were analysed by UMAP identifying four major continents and several smaller islands (Figure 1A). The relative expression of each marker on the UMAP (Supplementary Figure 3) revealed that the main continent on the left represents cytotoxic (CD8^+^) T-cells together with double negative (CD4^-^CD8^-^) T-cells. On the other hand, the main continent on the right is composed of helper (CD4^+^) T-cells. Likewise, the island on the bottom is mainly composed of monocytes, basophils and myeloid antigen presenting cells (APC), while the islands in the middle represent NK cells and Lδ T-cells. B-cells are represented in the top island together with dendritic cells.

To further refine the analysis, FlowSOM algorithm was applied to find similar cell subsets and separate them into metaclusters in an unsupervised manner (Figure 1B). The overlay of the FlowsSOM clustering on the UMAP representation (Figure1C) allowed us to perform a more exhaustive analysis, identifying a total of 73 clusters according to surface marker expression as shown in the heatmap (Figure 1D). Supplementary Table 2 shows an in-depth characterization of the phenotype of all clusters, which allowed the identification of 70 of them, since clusters 13, 32 and 33 could not be clearly identified. Finally, all clusters were further uploaded into the UMAP (Figure 1E) to determine not only how they relate to each other, but also to display their pseudoevolution.

**Figure 1.**
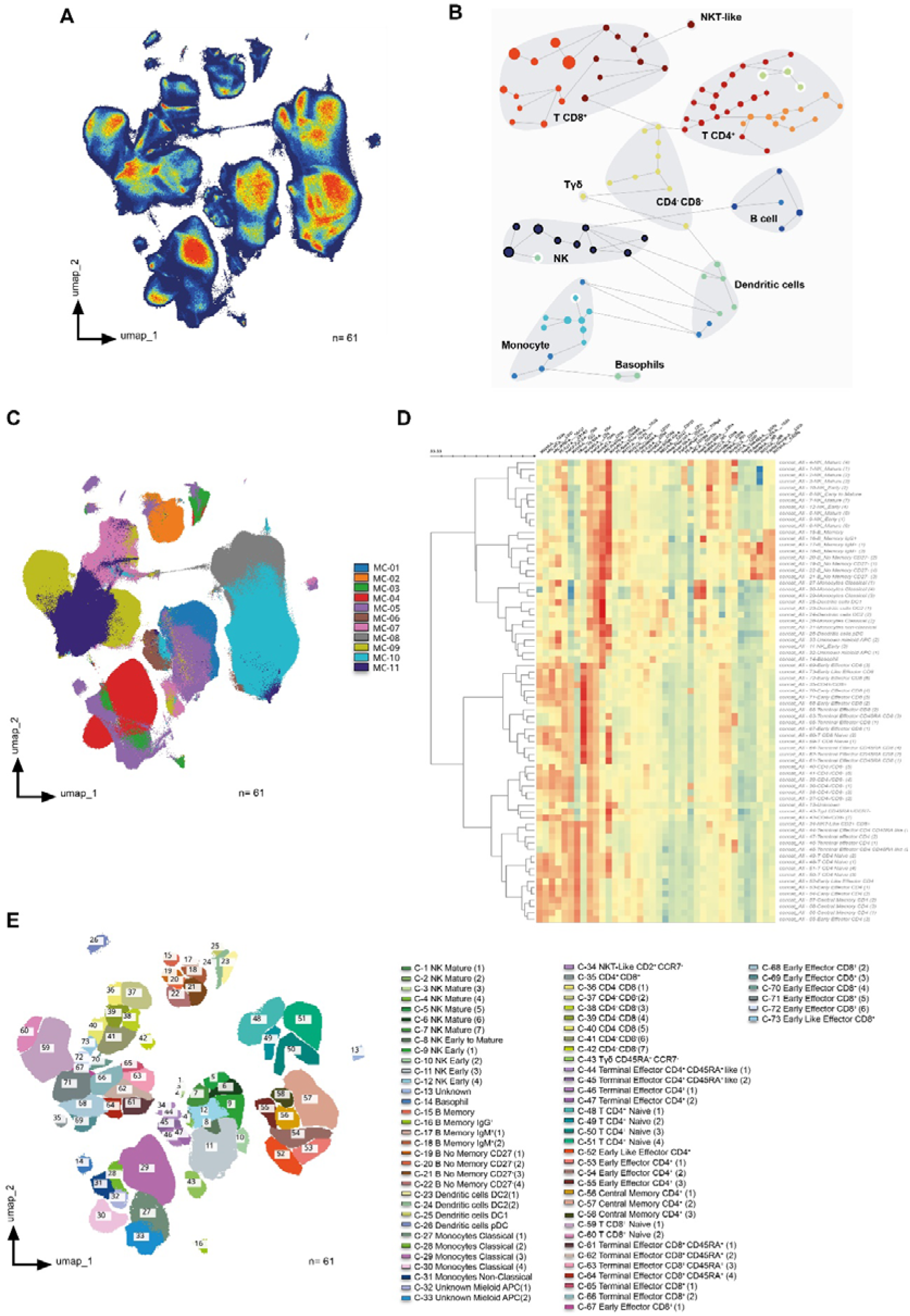
High dimensional analysis of peripheral blood mononuclear cells from controls and EoE patients. **(A)** UMAP density analysis representation performed on singlets corresponding to total viable circulating CD45^+^ cells from all samples. Samples were obtained from 19 controls and 24 patients at disease onset, as well as 9 responders and 9 non-responders to PPI therapy after 8-week treatment (a total of 61 samples). **(B)** FlowSOM clustering on total viable singlet CD45^+^ cells identified the main metaclusters on dataset: B-cells, NK cells, γδ T-cells, CD4^+^ T-cells, CD8^+^ T-cells, dendritic cells, basophils, monocytes, CD4^-^CD8^-^ T-cells, and NKT-like cells. **(C)** UMAP representation of all samples after non-supervised FlowSOM clusterization **(D)** Heatmap displaying the relative expression of each marker within each of the 73 identified clusters. Euclidean distance between clusters was calculated and represented by the dendrogram at the left side of the plot. **(E)** All 73 identified clusters were overlaid on the UMAP projection. Each identified cluster is tagged by a specific colour and number as shown in the legend.

### Peripheral immune profile differs between EoE patients and controls at baseline visit

After characterizing the different clusters, or immune subsets, present in our samples, we next addressed the immune differences found between controls and EoE patients at baseline visit. Volcano plot representation (Figure 2A) showed a significant deficit of plasmacytoid dendritic cells (pDC) (C-26) in EoE patients and an expansion of CD4^-^ CD8^-^ and early effector CD8^+^ T-cells (Clusters 37 and 69) at the time of disease diagnosis (Figure 2B). In order to further confirm these findings, classical gating strategies were applied as shown in Supplementary Figure 2, hence confirming that EoE patients display a deficit of circulating pDC at disease onset (Figure 2C).

**Figure 2.**
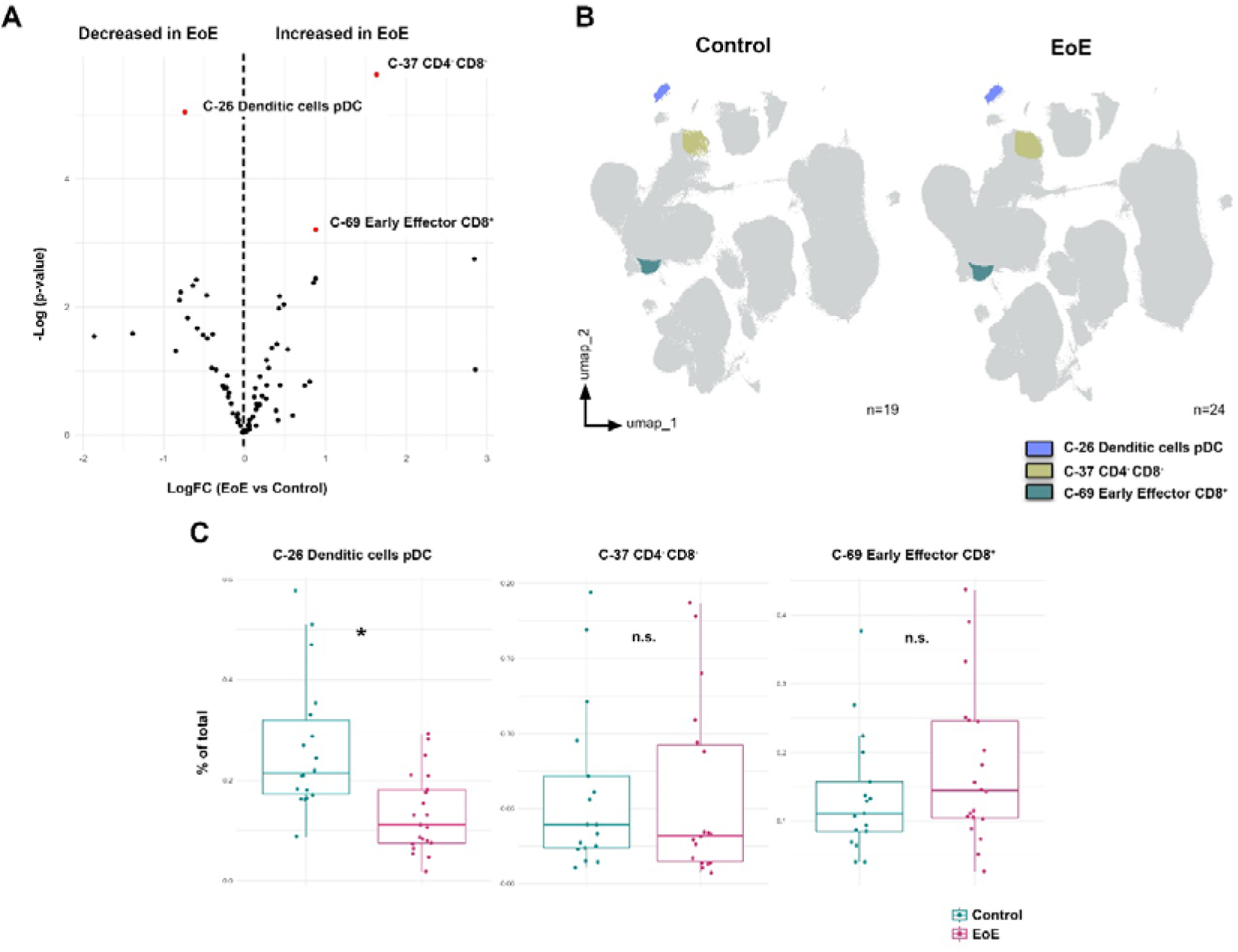
Peripheral immune profile differs between EoE patients and controls at baseline visit. **(A)** Volcano plot analysis comparing clusters from controls (n=19) and patients with eosinophilic esophagitis at disease diagnosis (EoE, n=24). LogFC and -Log(p-value) are shown. Clusters considered statistically significant are shown in red together with their nature as elucidated from Supplementary Table 2. **(B)** These differentially expressed clusters between control and EoE patients are further shown in the UMAP representation. **(C)** Validation of these clusters was performed following classical gating approaches (Supplementary Figure 1 and 2). *t*-test was applied in panel D, considering a p-value <0.05 as statistically significant (*p<0.05). Percent of total always refers to percentage of cells of the specified population relative to total PBMCs.

### Circulating pDC and classical monocyte levels at baseline are associated to PPI response

After describing that 3 clusters were differentially expressed on EoE patients at disease diagnosis, with a specific reduction of circulating pDCs as confirmed by classical gating approaches, we next aimed to address whether we could also identify specific clusters that might predict patient’s response to PPI treatment.

To that end, we compared the immune profile of responding (R) and non-responding (NR) patients (Table1) at disease onset (i.e., before PPI treatment [PrePPI]) (Figure 3A). Our results revealed that responding patients at diagnosis had higher levels of circulating pDC, myeloid antigen presenting cells and classical monocytes; together with lower levels of mature NK cells, and no memory B-cells and central memory CD4^+^ T-cells (Figures 3B and 3C). Of note, further analysis following classical gating approaches confirmed that PPI-responding patients had higher levels of circulating pDC and classical monocytes at disease onset compared to non-responders (Figure 3D).

**Figure 3.**
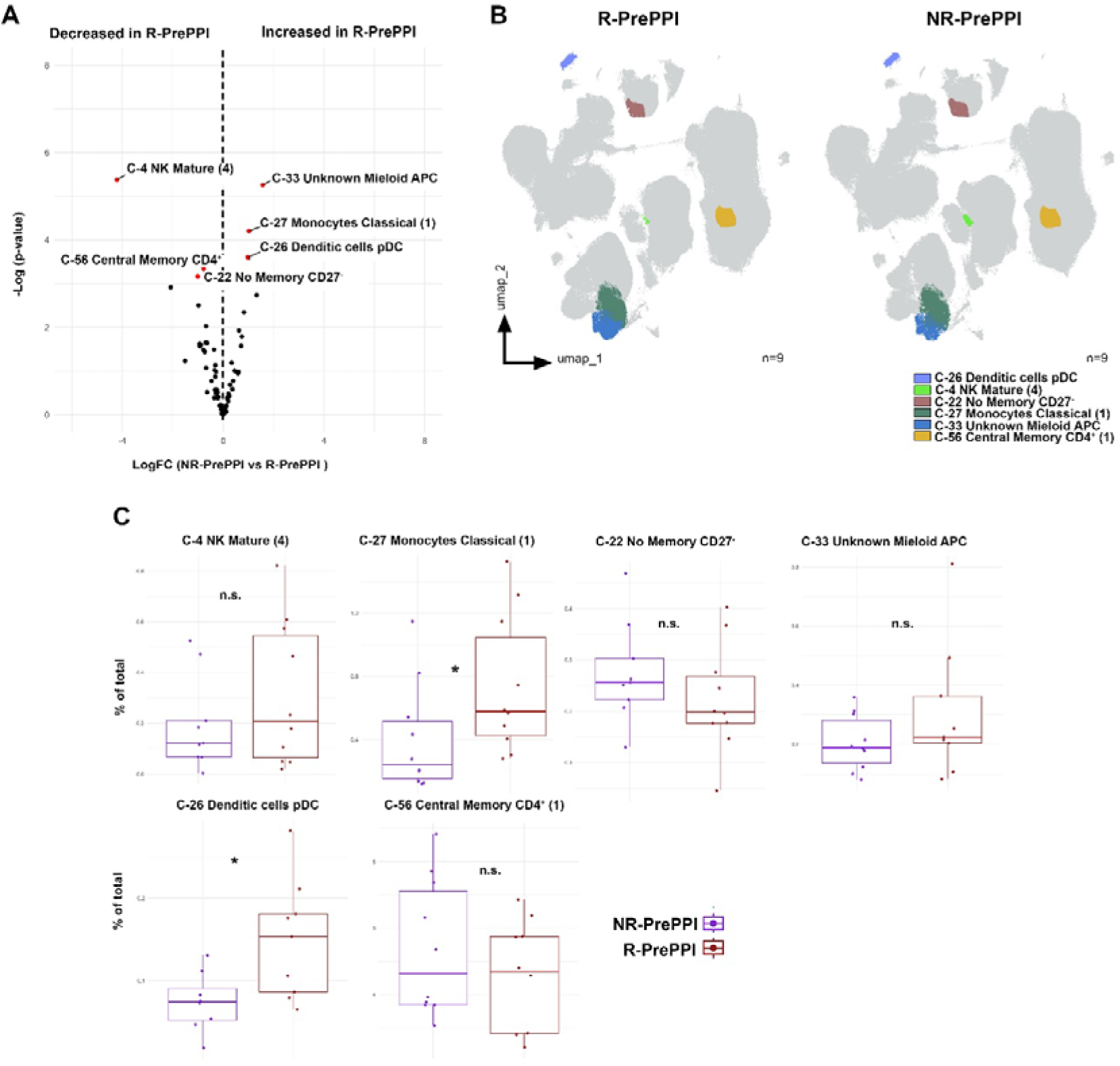
Circulating pDC and classical monocyte levels are associated to PPI response at disease diagnosis. **(A)** Volcano plot of differential analysis between responding (R, n=9) and non-responding patients (NR, n=9). LogFC and -Log(p-value) at baseline are shown. Clusters considered statistically significant are highlighted in red, showing their name and number. **(B)** UMAP representation from R and NR patients, in which significant clusters are coloured. **(C)** Validation by classic gating (Supplementary Figure 1 and 2) of the significant clusters previously defined. Boxplots of significant clusters represent individual percentage of total value, group medians as well as minimum and maximum values. Differences were analysed by *t*-test considering p-values <0.05 as statistically significant (*p<0.05, **p< 0.01, ***p< 0.001, ****p<0.0001) (R n=8, NR n=7). Responder (R), Non-Responder (NR), before treatment (PrePPI). Percent of total always refers to percentage of cells of the specified population relative to total PBMCs.

### Circulating pDC levels are restored in PPI-responding patients following treatment

Since patients with PPI responsive and non-responsive EoE displayed different immune cell levels at the time of disease diagnosis, we next aimed to address whether we could also identify specific clusters modulated by PPI treatment. To that end, we next compared the profile of EoE responsive (R) patients both before (PrePPI) and after (PostPPI) PPI therapy (Table1).

Our results revealed that, following treatment, circulating levels of pDC (C-26) and basophils (C-14) were increased in PPI-responding patients (Figures 4A and 4B), something particularly relevant in the case of circulating pDC as that increase was further confirmed by classical gating approaches (Figure 4C).

Based on these observations, we also assessed the immune cell dynamics in non-responding patients (Supplementary Figure 4A). The pDC cluster was increased in these patients following treatment (Supplementary Figure 4B); however, that observation could not be confirmed following classical gating strategy (Supplementary Figure 4C). Therefore, taken all together our results suggest that clinical response to PPI-treatment is associated with an increase in circulating pDC levels.

**Figure 4.**
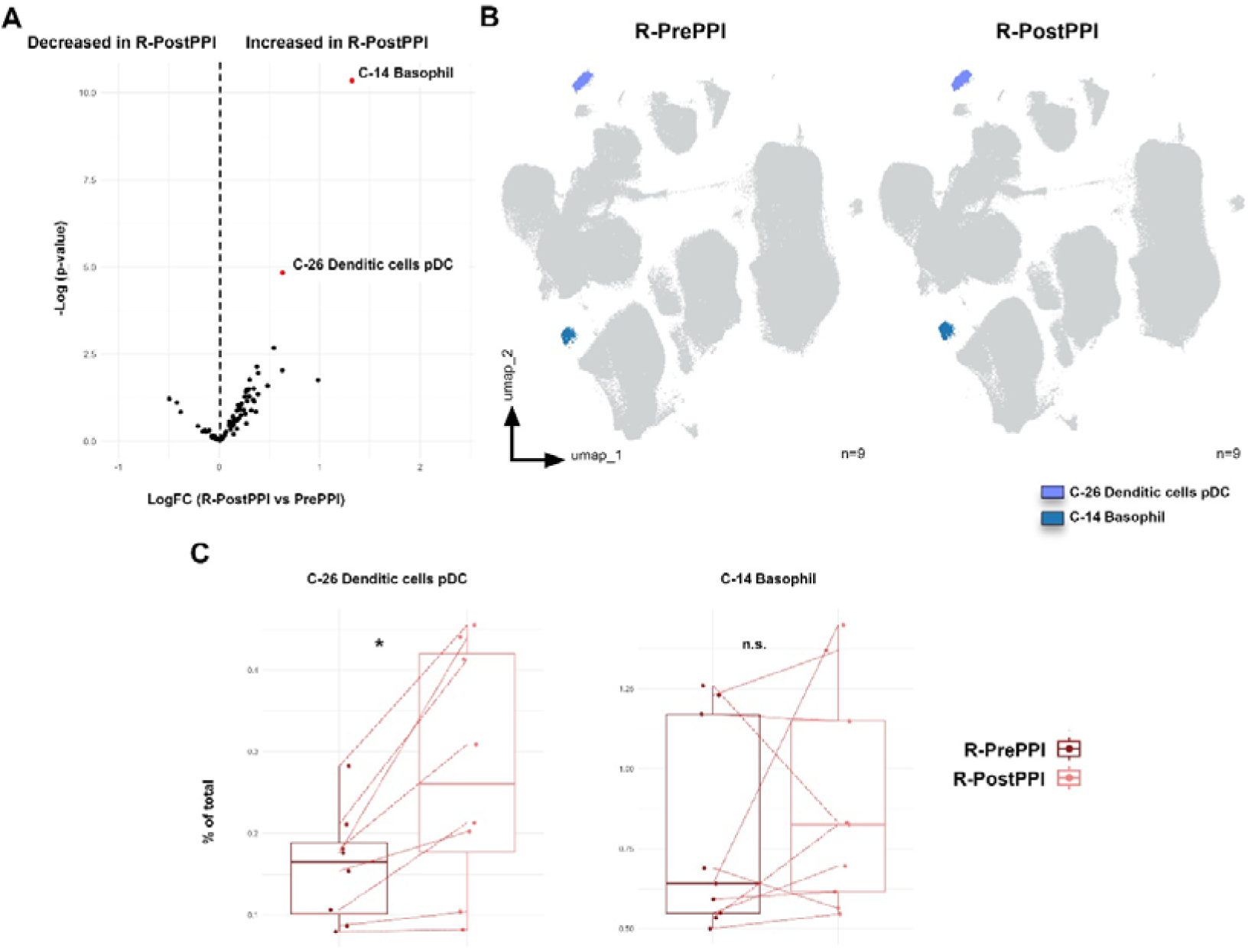
pDC and basophil levels are increased in responding patients upon PPI treatment. **(A)** Volcano plot analysis of the clusters comparing patients responding to proton pump inhibitor (PPI) treatment, both before (R-PrePPI) and after (R-PostPPI) therapy. LogFC and -Log(p-value) are shown. Clusters considered statistically significant are shown in red together with their nature as elucidated from Supplementary Table 2. **(B)** UMAP representation of these differentially expressed clusters in R-PrePPI and R-PostPPI. **(C)** Further validation of these clusters was performed following classical gating approaches as shown in Supplementary Figure 1 and 2. Boxplots of significant clusters represent individual percentage of total value, group medians as well as minimum and maximum values. Red line indicates paired PrePPI and PostPPI samples. Differences were analysed by paired *t*-test considering p-values <0.05 as statistically significant (*p<0.05, **p< 0.01, ***p< 0.001, ****p<0.0001) (n=8). Responder (R), before PPI treatment (PrePPI), after PPI treatment (PostPPI). Percent of total always refers to percentage of cells of the specified population relative to total PBMCs.

### Circulating pDCs are associated with EoE pathology and PPI-response

Having described that pDCs are differentially decreased in EoE patients at the moment of diagnosis, (Figure 2) while within EoE patients they are higher in those who respond to PPI treatment (Figure 3) and, indeed, they are further increased following such clinical intervention (Figure 4) although not in non-responding patients (Supplementary Figure 4), we next decided to focus on this cell population. Hence, further analysis confirmed that all EoE patients had lower levels of circulating pDCs at diagnosis, although these levels were lower in non-responding patients than in responders. Furthermore, in responders pDC levels were indeed further restored to control levels following treatment.

Since pDCs seem to be related to disease remission, we next hypothesized that cells might be correlated with the esophageal inflammatory state of the patient. To test this hypothesis, we studied the correlation between the maximum eosinophil count in the esophageal biopsy and pDC levels (Figure 5B). We found a negative correlation (r =-0.44 p-value= <0.001), suggesting the implication of pDCs in EoE pathogenesis.

**Figure 5.**
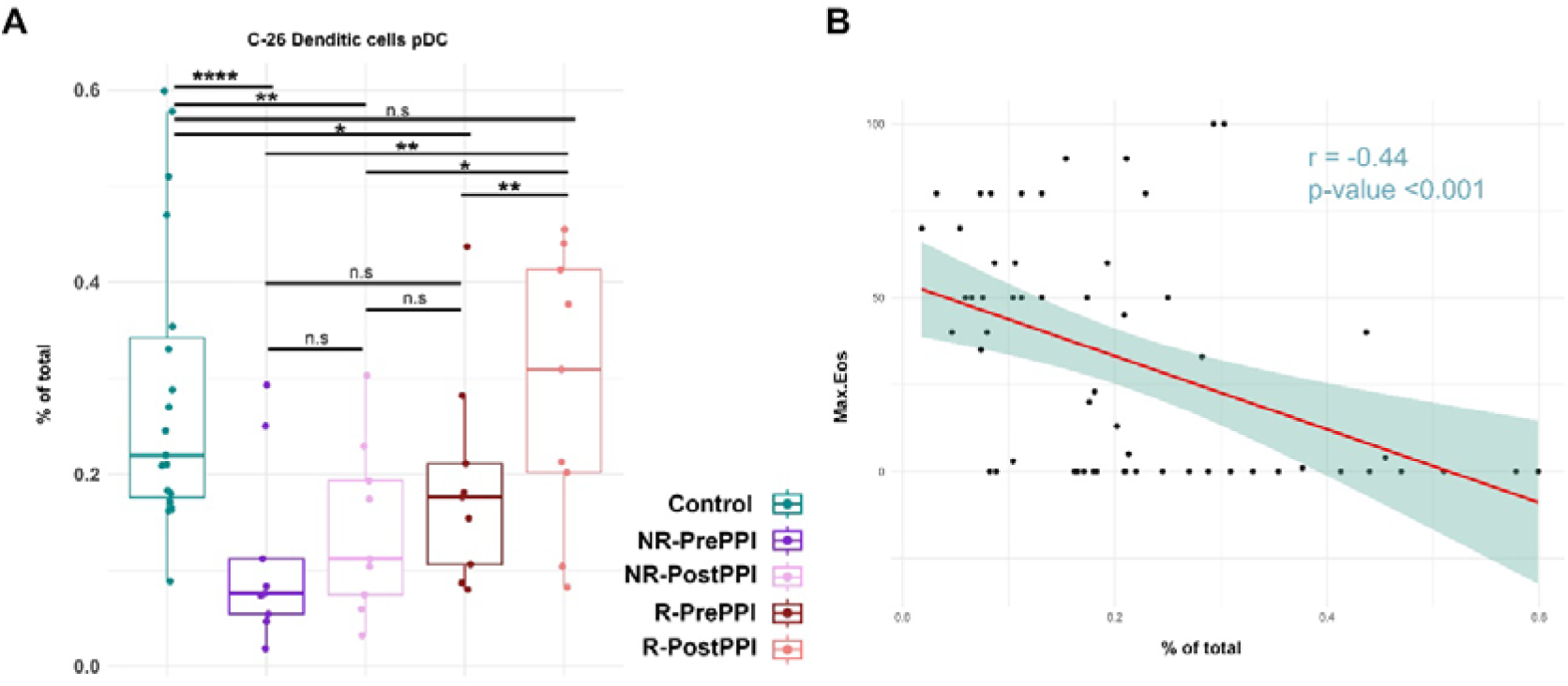
Circulating pDC are associated with EoE pathology and PPI-response. **(A)** Boxplots represent levels of pDC in control and EoE patients before and after PPI treatment, as individual percentages of total counts as well as group medians and minimum and maximum values. Differences were analysed by *t*-test, considering p-values <0.05 as statistically significant. **(B)** Pearson correlation between pDC and peak eosinophil count. Pearson correlation coefficient and p-value are shown. (Control n=19, R n=9, NR n=9). Responder (R), Non-Responder (NR), before PPI treatment (PrePPI), after PPI treatment (PostPPI) (*p<0.05, **p< 0.01, ***p< 0.001, ****p<0.0001).

### Differential activation profile of circulating pDCs in EoE and remission

Next, we assessed the expression of the chemokine receptors CCR7 and CXCR3 on circulating pDC, as they mediate pDC migration towards the lymph nodes (LN) and peripheral sites of inflammation^28,29^ (Figure 6A). CXCR3^+^ pDC (either CCR7^+^ or CCR7^-^) were increased in PPI responding patients following treatment (Figure 6B) suggesting a potential mechanism of action for pDC related with their infiltration in the esophagus.

**Figure 6.**
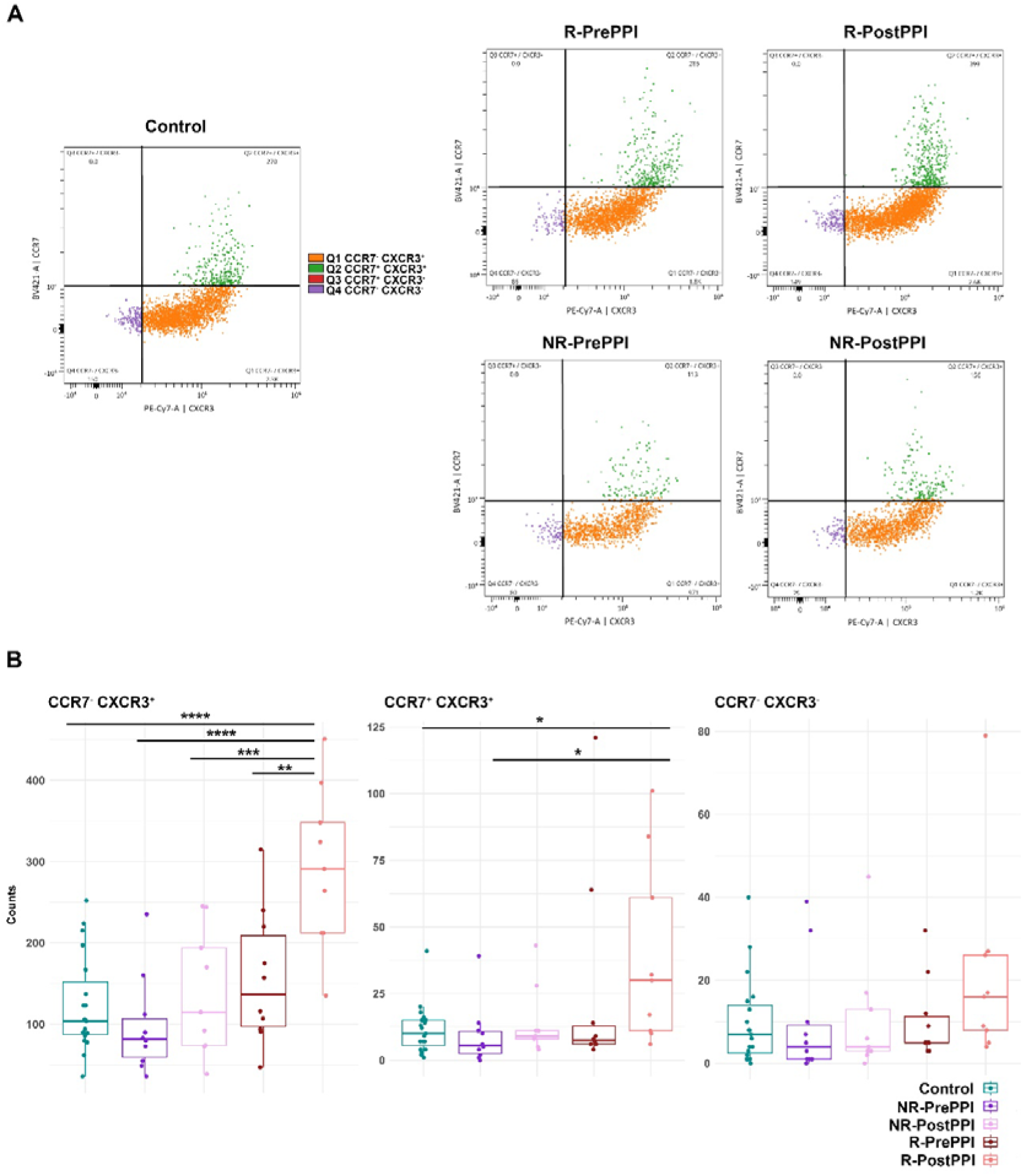
Differential activation profile of circulating pDC in EoE and remission. **(A)** Gating of activated pDC subpopulations according to their CCR7 and CXCR3 expression: CCR7^-^/CXCR3^+^(blue), CCR7^+^/CXCR3^+^ (orange), CCR7^+^/CXCR3^-^ (green), CCR7^-^/CXCR3^-^ (red). Dot plots show representative distribution of these subpopulations in each cohort. **(B)** Boxplot representation of the different pDC activation profiles in the five cohorts . Total count, group median and minimum and maximum values are represented. Differences were analysed by multiple comparison ANOVA followed by *post hoc* Fisher’s test, considering p-values <0.05 as statistically significant (*p<0.05, **p< 0.01, ***p< 0.001, ****p<0.0001). (Control n=19, R n=9, NR n=9). Responder (R), Non-Responder (NR), before treatment (PrePPI), after treatment (PostPPI).

### pDC infiltration in esophageal biopsies of EoE patients

Finally, after describing the activation profile of the circulating pDC population, we studied by immunofluorescence assay the infiltration of these cells in esophageal biopsies of 7 healthy controls and 13 EoE patients (6 R and 7 NR) before and after PPI treatment (Figure 7). pDCs were characterized as CD123^+^ (red) HLA-DR^+^ (green) CD11c ^-^ (blue) cells and quantified per mm^2^ of tissue (Figure 7A). The number of tissue pDCs was significantly higher in EoE patients than in controls (Figure 7B), being higher in NR-prePPI when compared to R-prePPI (Figure 7C). The number of tissue pDCs was reduced in NR and R patients after PPI treatment although differences were not statistically significant. However, R-postPPI values were the closest to those of controls (Figure 7C).

In general, tissue pDCs inversely correlated with the circulating pDC count and were directly related with the number of eosinophils per mm^2^ (Figure 7D,E). Thus, levels of this immune population seem to be related with PPI-response and EoE activity.

**Figure 7.**
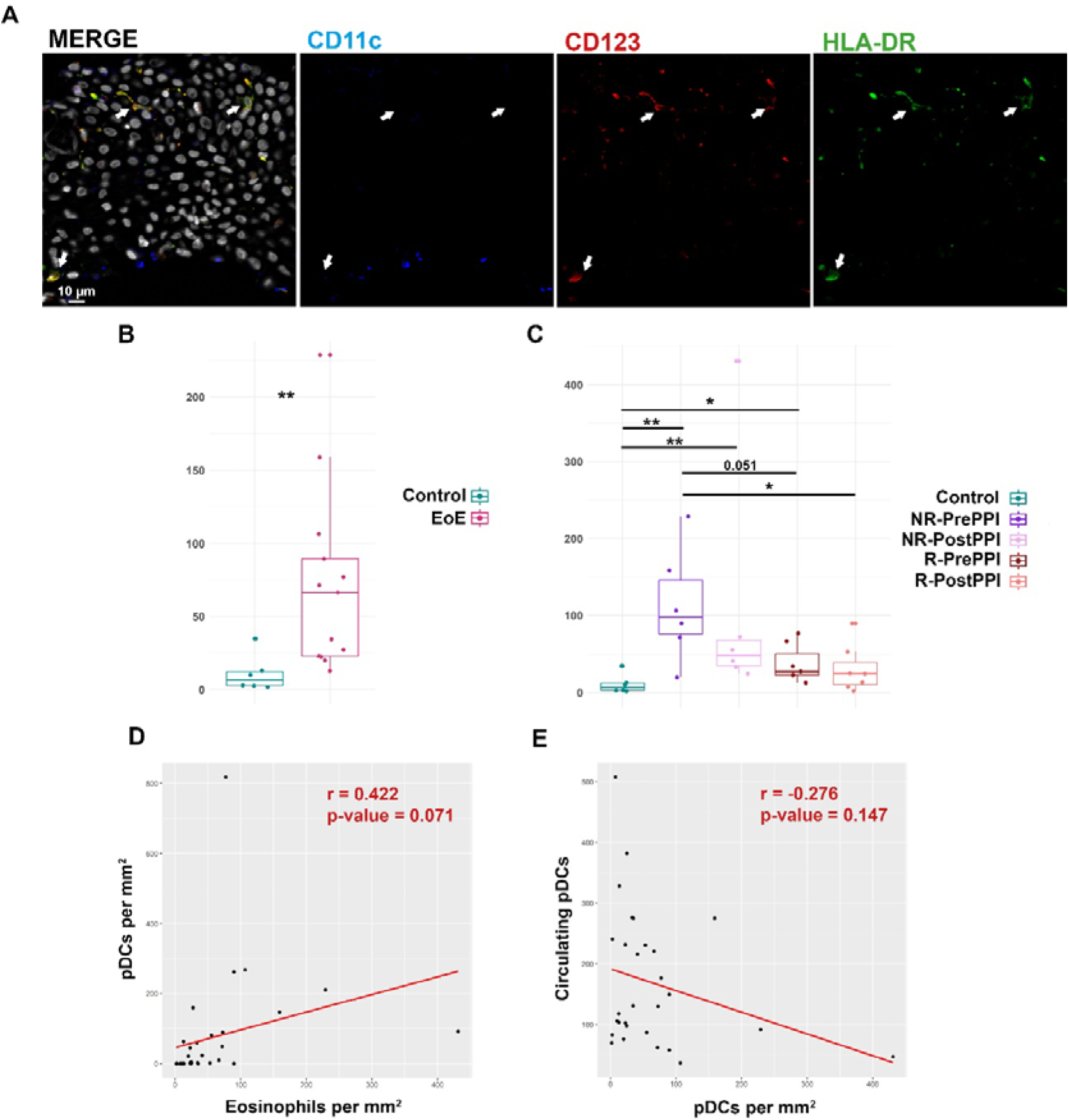
pDC infiltration in esophageal biopsies of EoE patients. **(A)** Immunofluorescence analysis of pDC infiltrate in esophageal biopsies from EoE patients. pDCs were visualized as CD123^+^(red) CD11c^-^ (blue), HLA-DR^+^ (green) cells. Arrows point to representative examples of pDC. Scale bar is shown. Nuclei were stained with DAPI (represented in grey in merged images). **(B)** Boxplot representation of infiltrating pDCs in controls (n=7) and EoE (n=13) patients at baseline. Relative count per mm^2^, group median and minimum and maximum values are represented. Differences were analysed by Mann-Whitney test, considering p-values <0.05 as statistically significant (*p<0.05, **p< 0.01, ***p< 0.001, ****p<0.0001). **(C)** Boxplot representation of infiltrating pDCs in controls (n=7), responder (n=6) and non-responder (n=7) pre-PPI and post-PPI. Relative count per mm^2^, group median and minimum and maximum values are represented. Differences were analysed by Mann-Whitney test, considering p-values <0.05 as statistically significant (*p<0.05, **p< 0.01, ***p< 0.001, ****p<0.0001). Responder (R), Non-Responder (NR), before treatment (PrePPI), after treatment (PostPPI). **(D)** Pearson correlation between tissue eosinophil count and tissue pDCs. Pearson correlation coefficient and p-value are shown. **(E)** Pearson correlation between circulating pDC and tissue pDCs. Pearson correlation coefficient and p-value are shown.

## DISCUSSION

Using state-of-the-art spectral cytometry, we hereby describe, for the first time to our knowledge, a unique fingerprint in the circulating immunome of patients with EoE at the time of disease diagnosis characterized by a specific deficit of circulating pDCs. Indeed, their levels were further decreased at diagnosis in patients who did not respond to PPI treatment, while among those who did, circulating levels of this cell subset were restored to normal levels upon treatment. Moreover, when we evaluated the presence of pDCs in the esophagus their numbers correlated with the eosinophilic count, showing both an inverse correlation with circulating pDCs. Hence, our findings suggest a central role of pDCs in the pathogenesis of EoE and reveal them as potential novel non-invasive biomarkers to aid on disease diagnosis and predict subsequent response to PPI therapy.

In addition to our novel findings on pDCs, we found an increase in CD4^-^CD8^-^ T-cells and CD8^+^ Early Effectors (C-37 and C-69, respectively) in EoE patients at diagnosis, although these results could not be further confirmed by classical gating approaches. Increased levels of T-cells could be related with the active role of CD8^+^T cells in the esophageal inflammation in EoE^30,31^ while the change in CD4^-^CD8^-^ T-cells might be associated with their cytotoxic activity as described for other pathologies ^32^, despite no role for these cells has been found in EoE.

PPI therapy is widely used in the management of EoE^16,33^, with a histologic remission rate of approximately 50%^16^. The reduction in Th2 signalling produced by this therapy leads to an improvement in the structural characteristics of the esophagus, accompanied by decreased eosinophil count. In our study, non-responding patients presented significant differences from responding patients (Figure 3A). When we studied these two groups PrePPI intake, non-responding patients had lower levels of pDCs, classical monocytes and an unknown myeloid APC cluster, but higher levels of non-memory B cells, mature NK cells and central memory CD4^+^ T-cells. These results might indicate an increased antigen presentation activity, since they have decreased numbers of circulating APC populations, which are key in homeostasis and allergy response^34^. Indeed, latest results from our group^35^ reinforce this hypothesis showing that non-responding patients’ esophageal proteomic profile when compared to responding patients (both PrePPI), have increased levels of proteins associated with antigen presentation. These patients might have a more altered barrier in the esophagus, which increases the risk of antigen exposure, thereby favouring EoE worsening, as described before^36–38^. Although these findings need a deeper characterization, they unveil for the first time a differential immunological profile between patients that will or will not respond to PPI therapy and could open the door to a better profiling of refractory patients.

In addition, when we studied immune dynamics during PPI treatment, PPI-responding patients displayed a significant increase in circulating basophils (C-14) and pDCs (C-26) (Figure 4A). Basophils have a pivotal role in atopic diseases and are a major source of Th2 immunomodulation. The increased levels of these cells in patients who achieve remission after PPI therapy could be related to the reestablishment of immunological homeostasis, since during inflammation basophils are able to penetrate the inflamed esophagus ^39^. A similar role may be played by circulating pDCs, which recover control values in PPI-responding patients PostPPI (Figure 5A). pDCs produce high levels of IFN-αβ (type I). This cytokine impairs Th2 responses by blocking the normal development of Th2 cells and production of Th2 cytokines (especially IL-5)^40,41^. The reduction in circulating pDCs during active esophageal inflammation could be indicating migration of these cells to the inflamed tissue to control the inflammatory response. Conceivably, circulating levels of these cells are restored when the patient achieves remission.

Importantly, the chemokine receptors CXCR3 and CCR7 related to pDC migration to tissues and homing to the LN^28,29,42^ were expressed in circulating pDCs (Figure 6A) in our cohorts. Indeed, responders presented the most activated pDCs after PPI treatment (Figure 5B) while no significant changes were found in the case of non-responding patients. Together with the previous results, the observed low levels of pDCs in PrePPI and PostPPI non-responding patients (Figure 5A) and their less active circulating profile (Figure 6B), could be linked to a poor inhibition of the Th2 response by type I IFN or/and a more exacerbated mucosal barrier alteration and therefore a worse prognosis^36,40^.

Focusing on promising minimally invasive biomarkers, pDCs are a potentially interesting candidate since they are highly related with EoE onset and the response to treatment. We have observed an inverse correlation between circulating pDCs and the peak eosinophil count in esophageal biopsies (Figure 5B). Moreover, when we evaluated tissue pDC levels in patient’s biopsies we found a positive correlation between these cells and the proportion of eosinophils per mm^2^ (Figure 7D), being also inversely correlated to pDC circulating levels (Figure 7E). In patients, the number of tissue pDCs was higher when the esophageal inflammation was active, reducing their abundance when the inflammation was under control (Figure7C). However, Responder patients do not reach control values after treatment despite pDC levels tend to be lower. This could be due to a non-complete recovery state, in which these cells participate in the restoration of the esophageal homeostasis as happens in other pathologies^43^ where tissue healing plays a key role. Thus, it may be that the evaluation of tissue pDC levels in long-term recovered patients would reveal closer numbers to the control.

Hence, this population is related with the inflammatory status of the esophagus, thus providing information of clinical diagnostic parameters that until now can only be measured through invasive procedures. In peripheral blood, previous works have related an increase in Th2 profile to active EoE^44^, while others have associated this to eosinophil phenotype or maturation state^45,46^. Also, single cell studies have described T-cell heterogeneity in the inflamed tissue, defining a specific enrichment in resident CD4^+^ T regulatory and Th2 effector cells together with an increased CD4^+^/CD8^+^ circulating T-cell ratio ^47^. Nevertheless, none of these parameters is being used as biomarker for EoE diagnosis, or for the prediction of response to PPI treatment, for which the peripheral eosinophil count has been proposed as a possible predictor^48^.

We are aware that advances in biomarker discovery in EoE are hampered by several pitfalls, such as the common concurrence of atopic diseases and the dissociation between the esophageal inflammation and patient’s symptoms^23^. Accordingly, despite our cohorts are highly controlled and paired samples from the same patients were analysed, further studies and validations are needed to confirm the utility of circulating pDCs as biomarker. The activity of this cluster must be compared in cohorts with other allergic and atopic conditions, since in these pathologies a decreased peripheral pDC count was found, together with pDC infiltration in the inflamed tissue^49–51^. Probably, its visualization through classic cytometry approaches and its combination with other non-invasive clinical parameters would be helpful to establish a specific signature for EoE management and PPI response prediction, also opening the door to study its utility in the case of other therapy options for EoE.

In summary, we have described, for the first time to our knowledge, the circulating immunome of EoE patients at the time of diagnosis highlighting as well the differences between PPI-responding and non-responding patients. Altogether, our study provides new insights on EoE immunity and sheds light on the characterization of this disorder, proposing a potential biomarker for diagnosis and prediction of response to treatment, which could improve decisions on better treatment options.

## Data Availability

All data produced in the present study are available upon reasonable request to the authors

## AUTHOR CONTRIBUTIONS

Conceived and designed the study: AJL, DB, CS, PM.

Participated in the clinical management of patients: SC, MTF-P, VM-D, JF-P, CS.

Performed the experiments: LU-T, FM-J, JH-V, LA-G, PM.

Analysed and discussed the data: LU-T, FM-J, JH-V, CR-R, DB, PM.

Wrote the paper: LU-T, DB, JH-V, CS, PM.

All the authors read, provided comments, and approved the final version of the manuscript.

## ABBREVIATIONS

EoE: Eosinophilic esophagitis
EoEHSS: Eosinophilic Esophagitis Histologic Scoring System
EREFS: EoE endoscopic reference score
GERD: Gastro-esophageal reflux disease
LN: Lymph node
LogFC: Log2 Fold Change
NR: Non-responder
PBMCs: Peripheral blood mononuclear cells
pDC: Plasmacytoid dendritic cells
PPI: Proton Pump Inhibitors
R: Responder
Th2: T helper 2 cells
UMAP: Uniform Manifold Approximation and Projection

## ACKNOWLEDGMENTS

We acknowledge, Instituto de Salud Carlos III, FEDER organization, Spanish Gastroenterology Association and Spanish Ministry of Science for the support to this study. Also, we would like to acknowledge the support from European Social Fund, NextGenerationEU, and Recovery, Transformation and Resilience Plan. We express our gratitude to Dr. Manuel Gómez for critical review of the manuscript and English editing.

