## Supplementary Files for "Circulating immunome fingerprint in eosinophilic esophagitis is associated with clinical response to proton pump inhibitor treatment"

### Supplementary Figure 1

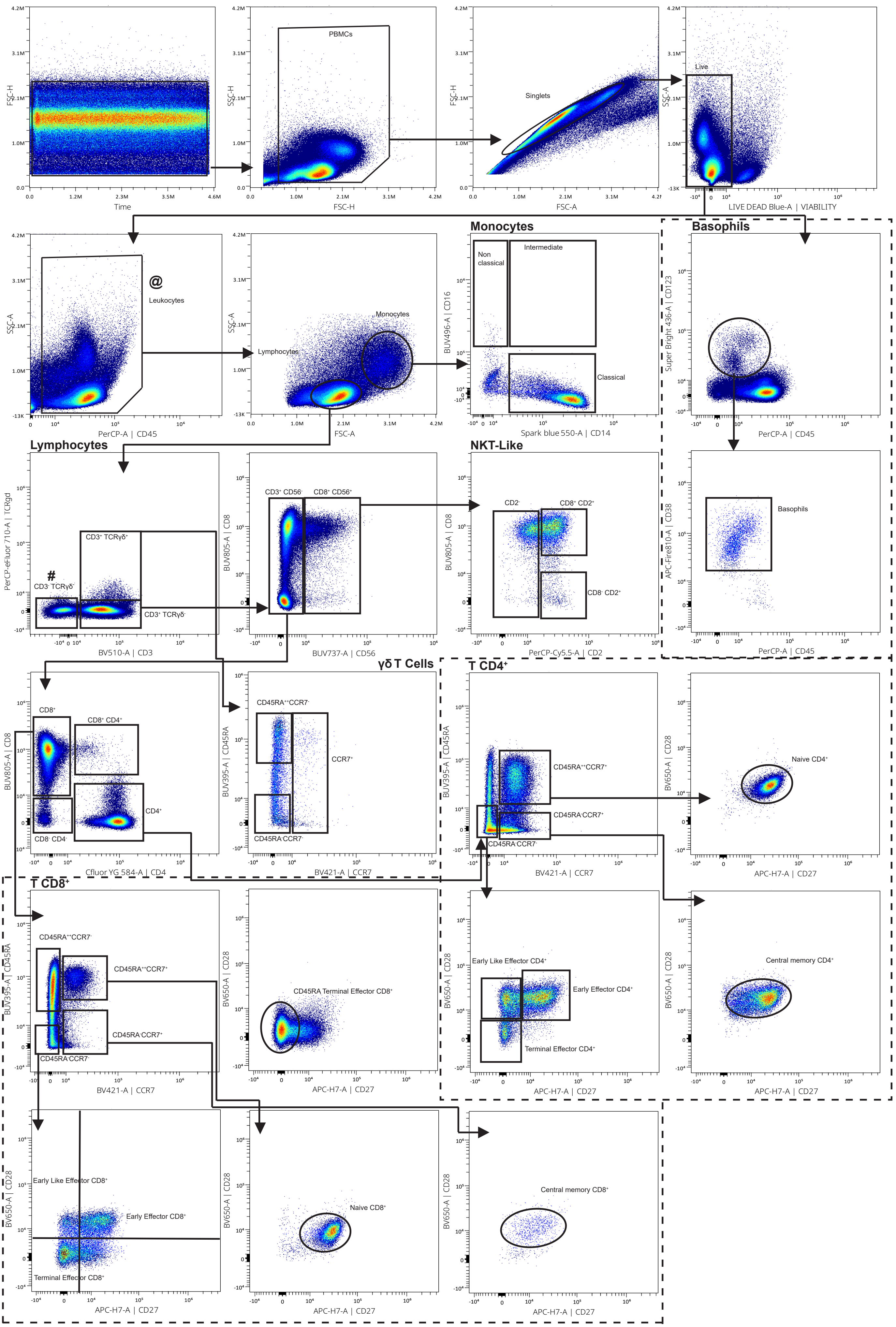

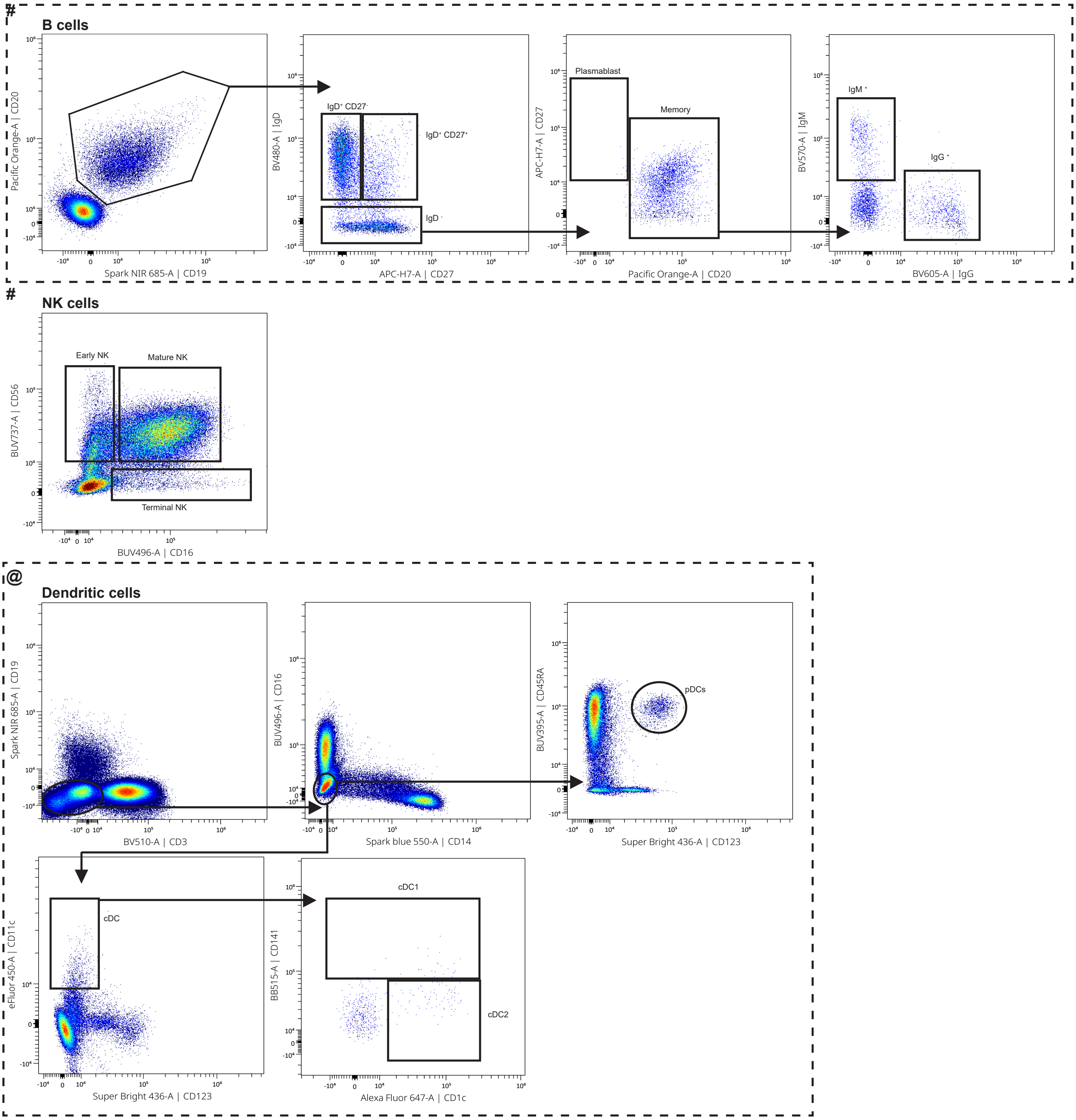


**Supplementary Figure 1. Hierarchical gating strategy for cluster classification.**
Representative gating strategy used to identify within total peripheral blood mononuclear cells (PBMC) the main leukocyte populations. Arrows and gates are used to visualize the relationships across plots. After doublets and dead cells were excluded, basophils were identified as $CD45^+CD123^+CD38^+$. Monocytes were gated based on FSC-A/SSC-A properties and further classified into non-classical ($CD14^-CD16^+$), intermediate ($CD14^+CD16^+$/low), and classical ($CD14^+CD16^-$). From the lymphogate, T-cell were identified based on the expression of CD3. Total Tγδ cells were identified as $CD3^+TCR\gamma\delta^+$ and subdivided based on the expression of CD45RA and CCR7. Total NKT-like cells were identified in the $CD3^+TCR\gamma\delta^-$ compartment as $CD56^+$. The inclusion of CD2 and CD8 further classified the NKT-like cells. Total T-cells were defined as $CD3^+TCR\gamma\delta^-CD56^-$ and further divided into $CD4^+$, $CD8^+$, $CD4^+CD8^+$ and $CD4^-CD8^-$ T-cells. Within total $CD4^+$ T-cells and $CD8^+$ T-cells, CCR7, CD45RA, CD27, and CD28 were further used to divide them into different T-cell phenotypes. B-cells were gated from the $CD3^-TCR\gamma\delta^-$ as $CD19^+$ and/or $CD20^+$ cells. B-cells were further classified as $IgD^+CD27^-$, $IgD^+CD27^+$, or $IgD^-CD27^{+/-}$; the $IgD^-CD27^{+/-}$ subset was divided into plasmablasts or $IgD^-$ memory B cells based on CD20 expression and CD27. Memory cells were classified in $IgM^+$ or $IgG^+$. NK cells were defined within the lymphogate as $CD3^-TCR\gamma\delta^-$ and classified as early NK ($CD56^+CD16^-$), mature NK ($CD56^+CD16^+$), and terminal NK ($CD56^-CD16^+$) cells. Dendritic cells were identified within $CD3^-CD19^-$ as $CD14^-CD16^-$. Further gating with $CD123^+CD45RA^+$ determined plasmacytoid DCs, (pDC) and $CD123^-CD11c^+$ as classical or conventional DC (cDC). cDC were divided into type 1 ($CD141^+$, cDC1) and type 2 ($CD1c^+$, cDC2).

### Supplementary Figure 2

##### C-56 Central Memory CD4⁺ (1)

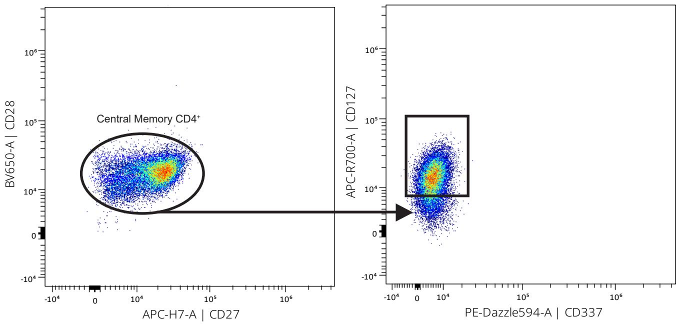


##### C-69 Early Effector CD8⁺

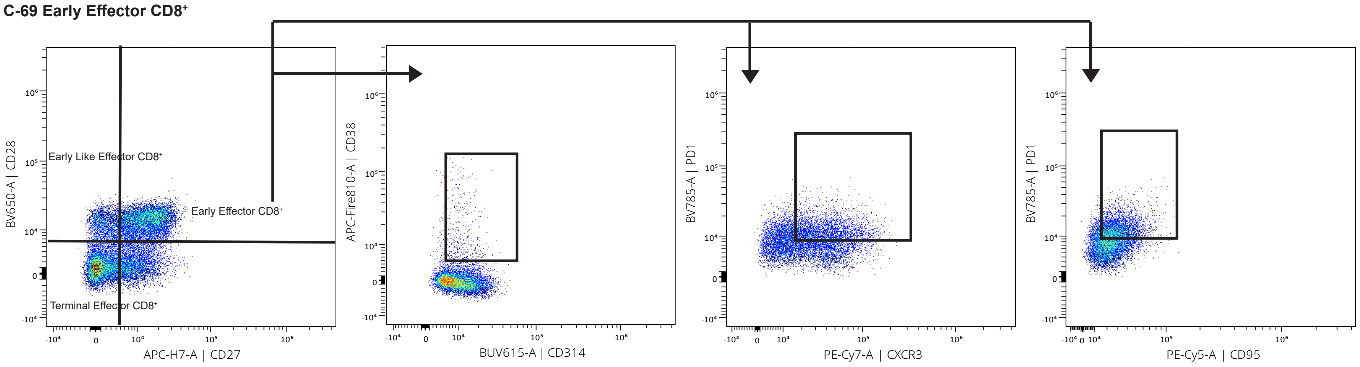


### C-37 CD4⁻ CD8⁻

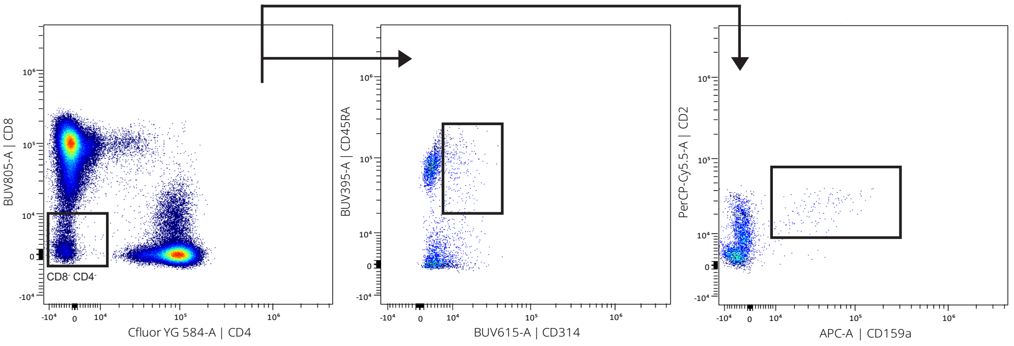


##### C-4 NK Mature (4)

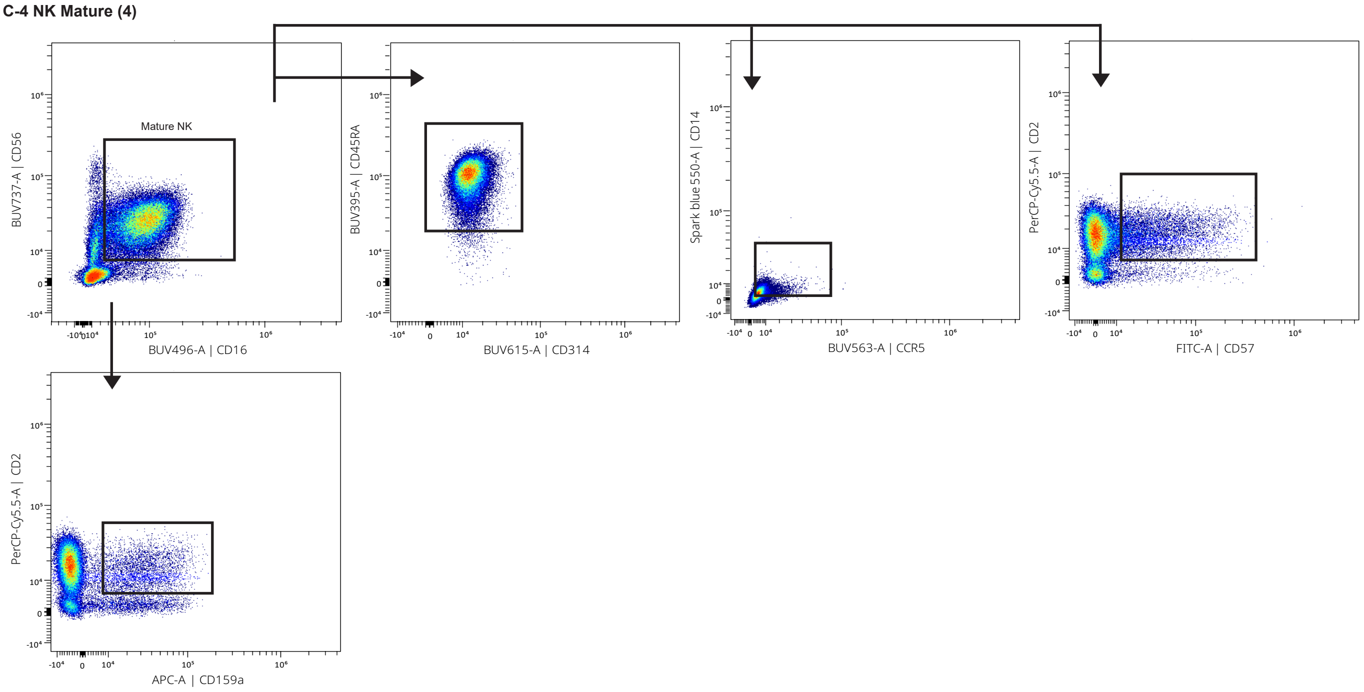

**C-22 No Memory CD27⁻**

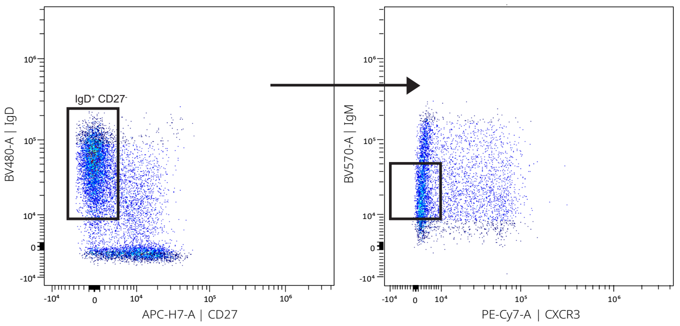


**C-26 Denditic cells pDC**

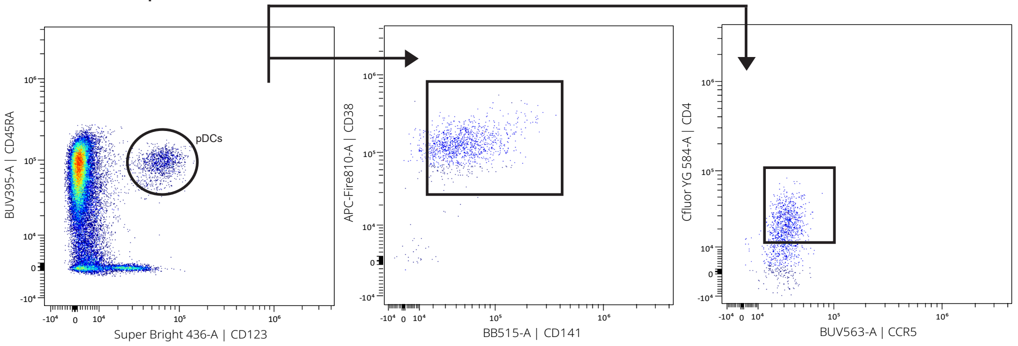


**C-27 Monocytes Classical (1)**

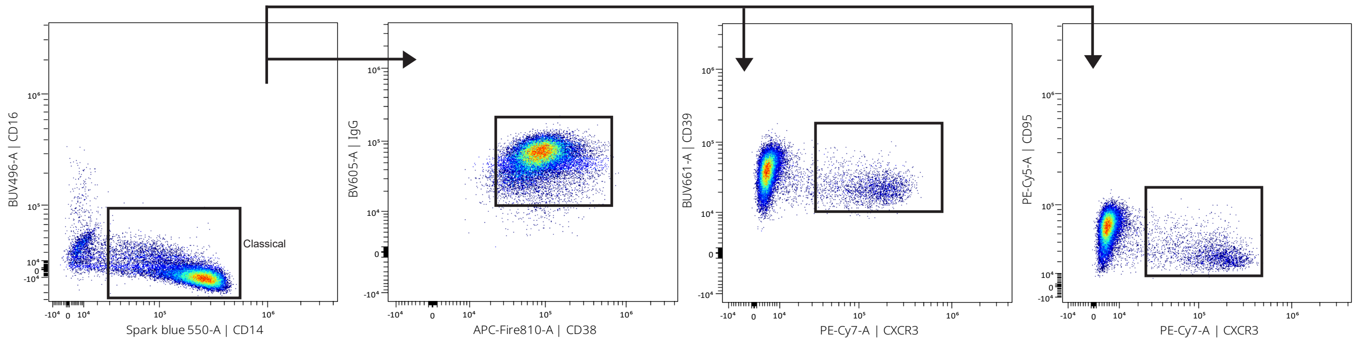


**C-33 Unknown Mieloid APC**

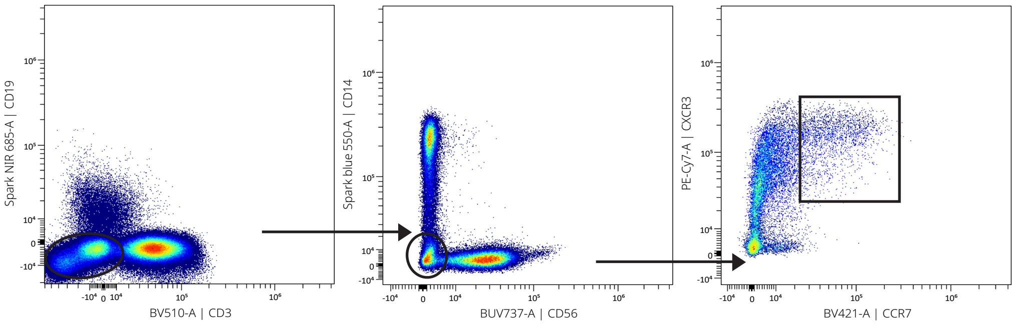


**C-14 Basophil**

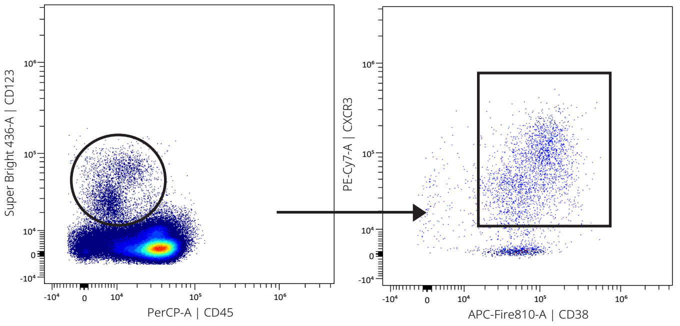

**Supplementary Figure 2. Gating strategy for significant cluster validation.**
Cell clusters identified in Table 1 which were statistically significant found among the different comparisons performed were validated further identified following classical hierarchical gating approaches. C-56 was gated from central memory CD4 CD127$^+$. C-69 was identified from early effector CD8 which were CD38$^+$ CD314$^+$ PD1$^+$ CXCR3$^+$CD95$^+$. C-37 was classified from CD8$^-$CD4$^-$ as CD314$^+$ CD45RA$^+$ CD2$^+$ CD159a$^+$. C-4 subset was identified as Mature NK CD45RA$^+$ CD314$^+$ CD14$^+$ CCR5$^+$ CD2$^+$ CD57$^+$ CD159a$^+$. C-22 was gated from No Memory CD27$^-$ with IgM$^+$ CXCR3$^-$. C-26 was defined as CD38$^+$ CD141$^+$ CD4$^+$ CCR5$^+$. C-27 were classified from classical monocytes IgG$^+$ CD38$^+$ CD39$^+$ CXCR3$^+$ CD95$^+$. C-33 was gated from CD19- CD3$^-$ CD14$^-$ CD56$^-$ CXCR3$^+$ CCR7$^+$. C-14 was defined as basophil CXCR3$^+$ CD38$^+$.

### Supplementary Figure 3

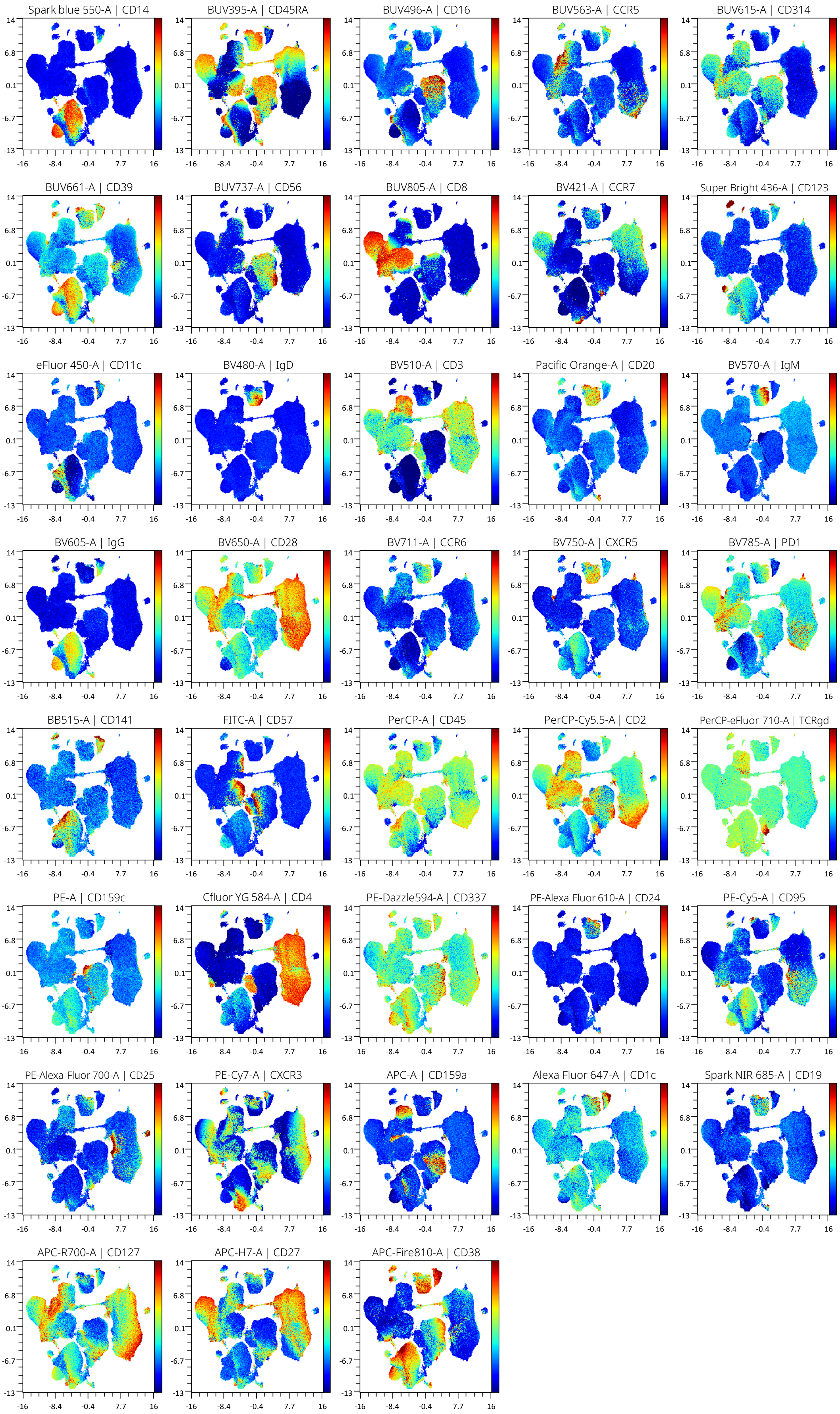

**Supplementary Figure 3. Marker expression on the UMAP from peripheral blood mononuclear cells.** Surface expression intensities of all 37 analysed markers is shown by a colour code based on the intensity. Red represents higher expression and blue represents lower expression.

#### Supplementary Figure 4

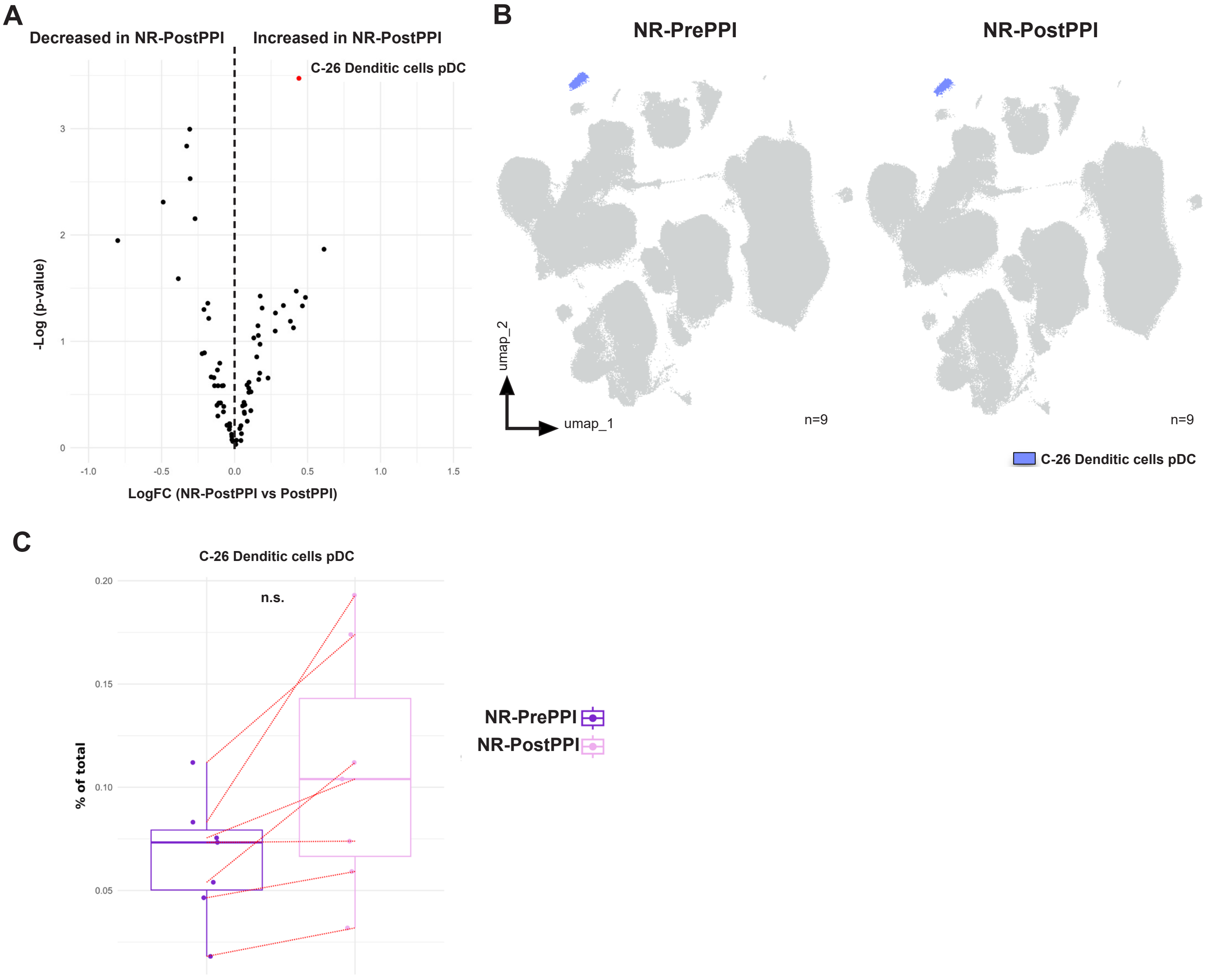


**Supplementary Figure 4. Non-Responders do not recover pDC values after treatment.** (A) Volcano plot of differential analysis between NR-PrePPI (n=9) and NR-PostPPI (n=9). LogFC and -Log(p-value) is represented. (B) UMAP representation from NR-PrePPI and PostPPI patients with MC-26 coloured. (C) Validation by classic gating of MC-26 as shown in Supplementary Figure 1 and 2. Boxplot with MC-26 where individual percent of total value, group median and minimum and maximum values is represented. Red line is indicating paired samples PrePPI and PostPPI. Statistical significance was established by paired t-test analysis considering a p-value <0.05 as statistically significant (*p<0.05) (n=7). Non-Responder (NR), before treatment (PrePPI), after treatment (PostPPI).

#### Supplementary Table 1

| Specificity | Fluorochrome | Clone | Provider |
|---|---|---|---|
| CCR5 | BUV563 | 2D7/CCR5 | BD Biosciences |
| CCR6 | BV711 | G034E3 | BioLegend |
| CCR7 | BV421 | G043H7 | BioLegend |
| CD1c | Alexa Fluor 647 | L161 | BioLegend |
| CD2 | PerCP-Cy5.5 | TS1/8 | BioLegend |
| CD3 | BV510 | SK7 | BioLegend |
| CD4 | cFluor YG584 | SK3 | CYTEK |
| CD8 | BUV805 | SK1 | BD Biosciences |
| CD11c | eFluor 450 | 3,9 | Thermo Fisher |
| CD14 | Spark Blue 550 | 63D3 | BioLegend |
| CD16 | BUV496 | 3G8 | BD Biosciences |
| CD19 | Spark NIR 685 | HIB19 | BioLegend |
| CD20 | Pacific Orange | HI47 | Thermo Fisher |
| CD24 | PE-Alexa Fluor 610 | SN3 | Thermo Fisher |
| CD25 | PE-Alexa Fluor 700 | CD25-3G10 | Thermo Fisher |
| CD27 | APC-H7 | M-T271 | BD Biosciences |
| CD28 | BV650 | CD28.2 | BioLegend |
| CD38 | APC-Fire 810 | HIT2 | BioLegend |
| CD39 | BUV661 | TU66 | BD Biosciences |
| CD45 | PerCP | 2D1 | BioLegend |
| CD45RA | BUV395 | 5H9 | BD Biosciences |
| CD56 | BUV737 | NCAM16.2 | BD Biosciences |
| CD57 | FITC | HNK-1 | BioLegend |
| CD95 | PE-Cy5 | DX2 | BioLegend |
| CD123 | Super Bright 436 | 6H6 | Thermo Fisher |
| CD127 | APC-R700 | HIL-7R-M21 | BD Biosciences |
| CD141 | BB515 | 1A4 | BD Biosciences |
| CD159a | APC | REA110 | Miltenyi |
| CD159c | PE | REA205 | Miltenyi |
| CD314 | BUV615 | 1D11 | BD Biosciences |
| CD337 | PE-Dazzle 594 | P30-15 | BioLegend |
| CXCR3 | PE-Cy7 | G025H7 | BioLegend |
| CXCR5 | BV750 | RF8B2 | BD Biosciences |
| IgD | BV480 | IA6-2 | BD Biosciences |
| IgG | BV605 | G18-145 | BD Biosciences |
| IgM | BV570 | MHM-88 | BioLegend |
| PD-1 | BV785 | EH12.2H7 | BioLegend |
| TCRγδ | PerCP-eFluor 710 | B1.1 | Thermo Fisher |
| Viability | Live Dead Blue-A | - | Thermo Fisher |

**Supplementary Table 1.**
Specificity, fluorochrome, clone and provider of the different antibodies used.

#### Supplementary Table 2

| C | Population | Subset | Phenotypic expression | Functional expression |
|---|---|---|---|---|
| 1 | NK | Mature (1) | **CD16** CD56 CD45RA | CD159c **CXCR3** CD314 |
| 2 | NK | Mature (2) | **CD16** CD56 CD45RA | **CD2 CD57** CD159c **CXCR3** CD314 |
| 3 | NK | Mature (3) | **CD16** CD56 CD45RA | CD2 CD57 CD159c CD314 |
| 4 | NK | Mature (4) | **CD16** CD56 CD45RA | CD314 **CD14 CCR5 CD2 CD159a CD57** |
| 5 | NK | Mature (5) | **CD16** CD56 CD45RA CD38 CD8 | **CD2** CD314 |
| 6 | NK | Mature (6) | **CD16** CD56 CD45RA | **CD2** CD314 |
| 7 | NK | Mature (7) | **CD16** CD56 CD45RA CD38 | CD314 |
| 8 | NK | Early to Mature | **CD16dim** CD56 CD45RA$^{++}$ | CD2 CD314 CD159a |
| 9 | NK | Early (1) | CD45RA CD56 CD38 | CD314 CD337 |
| 10 | NK | Early (2) | CD45RA CD56 CD38 | CD314 **CD159a** CD337 **CD2** |
| 11 | NK | Early (3) | CD45RA CD56 CD38 | **CXCR3** |
| 12 | NK | Early (4) | CD45RA CD56 | |
| 13 | Unknown | | CD45RA CD25 CD127 | |
| 14 | Basophil | | CD123 CD38 | CXCR3 |
| 15 | B | Memory | CD20 CD19 CD45RA CD27dim CD38 | CD39 CCR7 CCR6 CXCR5 CD95 CD1c |
| 16 | B | Memory IgG$^{+}$ | CD20 CD19 CD45RA$^{+}$ **IgG** CD38 | CCR6 CXCR5 CD159c CD337 |
| 17 | B | Memory IgM$^{+}$ (1) | CD20 CD19 CD45RA$^{++}$ **IgM** CD24 CD25 CD38 | CCR7 **CCR5** CCR6 CXCR5 PD1 **CD337** CD1c |
| 18 | B | Memory IgM+ (2) | CD20 CD19 CD45RA$^{++}$ **IgM** CD24 CD25 CD38 | CCR7 CCR6 CXCR5 PD1 CD1c |
| 19 | B | No Memory CD27$^{-}$ (1) | CD20 CD45RA CD38 | CCR7 CCR6 CXCR5 **CXCR3** |
| 20 | B | No Memory CD27$^{-}$ (2) | CD20 CD19 CD45RA$^{++}$ **IgD** CD38 | CCR7 CCR6 CXCR5 **CXCR3** |
| 21 | B | No Memory CD27$^{-}$ (3) | CD20 CD19 CD45RA$^{++}$ **IgD**$^{++}$ **IgMdim** CD38 | CCR7 CCR6 CXCR5 **PD1** |
| 22 | B | No Memory CD27$^{-}$ (4) | CD20 CD19 CD45RA IgD CD38 | CCR7 CCR6 CXCR5 |
| 23 | Dendritic cells | cDC2 (1) | **CD11c** CD141 CD38 CD1c | CD39 CCR5 CD95 |
| 24 | Dendritic cells | cDC2 (2) | **CD11c** CD141 CD38 CD1c | CD39 CCR5 CD95 CXCR3 |
| 25 | Dendritic cells | cDC1 | **CD11c** CD141 CD38 CD1c | CD39 CCR5 CD95 CXCR3 |
| 26 | Dendritic cells | pDC | **CD123 CD45RA** CD141lo CD38 CD4 | CCR5 CXCR3 |
| 27 | Monocytes | Classical (1) | **CD14**$^{++}$ IgG CD38 | CD39 **CXCR3** CD95 |
| 28 | Monocytes | Classical (2) | **CD14** CD45RA CD11c IgG CD141 CD38 | CD95 CD39 |
| 29 | Monocytes | Classical (3) | **CD14** IgG CD38 | CD95 CD39 |
| 30 | Monocyte | Classical (4) | CD14 CD39 IgG CD95 | |
| 31 | Monocytes | Non-classical | **CD16** CD45RA CD11c CD141 CD38 | |
| 32 | Unknown | myeloid antigen presenting cells (1) | CD45RAdim CD11c CD38 | |
| 33 | Unknown | myeloid antigen presenting cells (2) | CD123 CD11c CD141 CD38 | **CXCR3** |
| 34 | NKT-Like cells | CD2$^{+}$ CD8$^{+}$ | CD3 CD4 **CD8dim CD2** CD45RA CD56 | CD57 |
| 35 | Tcell | CD4$^{+}$ CD8$^{+}$ | CD3 **CD4 CD8** CD28 CD27 CD25 | CD2 CD95 |
| 36 | Tcell | CD4$^{-}$ CD8$^{-}$ (1) | CD3 CD28 CD27dim | CD314 **CCR5** CD2 **CD159a CXCR3** |
| 37 | Tcell | CD4$^{-}$ CD8$^{-}$ (2) | CD3 CD45RA | CD314 CD2 CD159a |
| 38 | Tcell | CD4$^{-}$ CD8$^{-}$ (3) | CD3 CD45RA CD27dim | CD314 CD2 |
| 39 | Tcell | CD4$^{-}$ CD8$^{-}$ (4) | CD3 CD28 CD27dim | CD314 **CCR5** CD2 **CXCR3** |
| 40 | Tcell | CD4$^{-}$ CD8$^{-}$ (5) | CD3 CD28 | **CCR5** CD2 |
| 41 | Tcell | CD4$^{-}$ CD8$^{-}$ (6) | CD3 CD28dim CD27dim | CCR5 **CXCR3** |
| 42 | Tcell | CD4$^{-}$ CD8$^{-}$ (7) | CD3 CD45RA CD27 CD28 | CXCR3 **CCR7** |
| 43 | Tcell | γδ CD45RA$^{+}$/CCR7$^{-}$ | CD3 **TCRγδ CD45RA** CD141 CD25 CD1c | CD314 CCR6 CD2 |
| 44 | Tcell | CD4 Terminal Effector CD45RA like (1) | CD3 CD4 **CD45RA** CD56dim | CD2 |
| 45 | Tcell | CD4 Terminal Effector CD45RA like (2) | CD3 CD4 **CD45RA** | CD2 **CD57** |
| 46 | T cell | Terminal effector CD4 (1) | CD3 CD4 | CD2 PD1 **CD57** |
| 47 | T cell | Terminal effector CD4 (2) | CD3 CD4 | CD2 PD1 |
| 48 | Tcell | CD4 Naive (1) | CD3 CD4 **CD45RA**$^{++}$ **CCR7** CD28 CD27 | |
| 49 | Tcell | CD4 Naive (2) | CD3 CD4 **CD45RA CCR7** CD28 CD27 | |
| 50 | Tcell | CD4 Naive (3) | CD3 CD4 **CD45RA CCR7** CD28 CD27 | CXCR3 |
| 51 | Tcell | CD4 Naive (4) | CD3 CD4 **CD45RA**$^{++}$ **CCR7** CD28 CD27 | CXCR3 |
| 52 | Tcell | Early like Effector CD4 | CD3 CD4 **CD28** CD127 CD25lo | CD2 CD95 |
| 53 | Tcell | Early Effector CD4 (1) | CD3 CD4 **CD28 CD27** CD127 | CD2 **CXCR3** CD95 |
| 54 | Tcell | Early Effector CD4 (2) | CD3 CD4 **CD28 CD27** CD127 | **CD39dim** CD2 CD95 |
| 55 | Tcell | Early Effector CD4 (3) | CD3 CD4 **CD28 CD27** CD25 | CD95 |
| 56 | Tcell | Central Memory CD4 (1) | CD3 CD4 **CCR7 CD28 CD27** CD127 | CD337 CD95 |
| 57 | Tcell | Central Memory CD4 (2) | CD3 CD4 **CCR7 CD28 CD27** CD127 | **CD337 CXCR3** CD95 |
| 58 | Tcell | Central Memory CD4 (3) | CD3 CD4 **CCR7 CD28 CD27** | **CXCR3** CD95 |
| 59 | Tcell | CD8 Naive (1) | CD3 CD8 **CD45RA CCR7 CD27** | CD314 **CXCR3** |
| 60 | Tcell | CD8 Naive (2) | CD3 CD8 **CD45RA CCR7 CD27** | CD314 |
| 61 | Tcell | Terminal Effector CD45RA CD8 (1) | CD3 CD8 **CD45RA** | CD314 |
| 62 | Tcell | Terminal Effector CD45RA CD8 (2) | CD3 CD8 **CD45RA** | CD314 **CD2** |
| 63 | Tcell | Terminal Effector CD45RA CD8 (3) | CD3 CD8 **CD45RA** | CD314 **CD2 CD57** |
| 64 | Tcell | Terminal Effector CD45RA CD8 (4) | CD3 CD8 **CD45RA** | CD314 **CD2 CXCR3** |
| 65 | Tcell | Terminal Effector CD8 (1) | CD3 CD8 | CD314 CD2 **CD57** |
| 66 | Tcell | Terminal Effector CD8 (2) | CD3 CD8 | CD314 CD2 **PD1** |
| 67 | Tcell | Early Effector CD8 (1) | CD3 CD8 CD45RAlo **CD28 CD27** CD127 | CD314 CD2 PD1 |
| 68 | Tcell | Early Effector CD8 (2) | CD3 CD8 **CD28 CD27** | CD314 **CXCR3** |
| 69 | Tcell | Early Effector CD8 (3) | CD3 CD8 CD28 CD27 CD38 | CD314 PD1 CXCR3 CD95 |
| 70 | Tcell | Early Effector CD8 (4) | CD3 CD8 **CD28 CD27** CD127 | CD314 **CCR5 CD2 CXCR3 CD95 PD1 CD159a** |
| 71 | Tcell | Early Effector CD8 (5) | CD3 CD8 **CD28 CD27** CD127 | CD314 **CD2 PD1 CXCR3 CD95** |
| 72 | Tcell | Early Effector CD8 (6) | CD3 CD8 **CD28 CD27** CD127 CD2 | CD314 **CCR5 PD1** |
| 73 | Tcell | Early like Effector CD8 | CD3 CD8 **CD28** CD127 | CD314 **CCR5 CD2** |

**Supplementary Table 2.** Cell cluster identification from control and EoE patients.
For each of the 73 identified FlowSOM clusters its ontogeny and subset is shown, together with the specific subset, phenotype and expression of functional markers. Markers highlighted in bold denote differential expression within the same population.
